# Genotype-specific communication profiles in 79,518 individuals with neurodevelopmental disorders

**DOI:** 10.64898/2026.02.03.26345484

**Authors:** Cristiane Hsu, Alina Ivaniuk, Andres Jimenez-Gomez, Tobias Brünger, Christian M. Boßelmann, M. Scott Perry, Chiara Phan, Ana Arenivas, Haley O. Oyler, Natasha N. Ludwig, Costin Leu, Dennis Lal

**Author notes:** **Correspondence to:** Costin Leu, PhD, Department of Neurology, McGovern Medical School, UTHealth Houston, 6431 Fannin St, Houston, TX 77030, US.

## Abstract

**Rationale:** Neurodevelopmental disorders (NDDs) are characterised by significant challenges in communication, social interaction, and adaptive function, often impacting quality of life. Previous studies support genetic influences on the communication abilities of individuals with NDD, but were either limited to single genetic conditions or to small cohorts with a limited selection of communication measures.

**Methods:** We analysed caregiver-reported communication abilities in 79,518 individuals with NDD from the Simons Searchlight and SPARK registries: 4,439 with a CNV-based or monogenic NDD and 75,079 with autism spectrum disorder (ASD) without a known genetic cause (idiopathic ASD) as controls. For analysis, we a priori selected 10 communication-related measures based on their availability in the study cohorts, coverage of distinct communication aspects, and their frequent use in neurodevelopmental phenotyping, yielding 177,328 data points across all study cohorts. The individuals in the Searchlight registry were divided into a *Discovery* cohort (the 15 most prevalent genetic NDD conditions) and a *Confirmation* cohort (all other genetic NDD conditions). A second *Confirmation* cohort was generated using all individuals with genetic ASD forms from the SPARK registry. We then tested each of the three case cohorts and each genetic condition represented in the *Discovery* cohort against the ASD control cohort. Developmental trajectories were assessed through testing of participants grouped by age at evaluation.

**Results:** Measure-level analyses demonstrated significant associations between genetic status and communication abilities, differences in communication abilities between classes of genetic variants (monogenic vs. CNV-based NDDs), and variability between specific genetic NDD conditions. CNV-based NDDs showed milder communication impairment, outperforming idiopathic ASD controls in 9/10 communication measures, whereas monogenic NDD conditions had more pervasive impairments, especially in verbal communication. Although impaired in verbal communication, five monogenic NDD conditions showed at least suggestive strengths in nonverbal and social communication relative to idiopathic ASD controls (*CSNK2A1*, *CTNNB1*, *SETBP1*, *MED13L*, and *PPP2R5D*), specifically in using gestures. Developmental trajectory analyses revealed *STXBP1* as the gene group at highest risk of developmental stagnation in communication abilities.

**Conclusions:** These findings underscore the potential of precision speech-language pathology (SLP) approaches tailored to the specific verbal and nonverbal communication strengths and weaknesses of genetic groups. We also provide evidence for measurable improvements and declines in communication abilities with age at the group level, highlighting the need for developmentally informed care. By integrating genetic insights into clinical practice, precision SLP approaches may enhance communication outcomes and developmental progress and improve quality of life for individuals with genetic NDDs.

## Introduction

Neurodevelopmental disorders (NDDs) encompass a wide range of childhood-onset impairments in communication, social interaction, and adaptive functioning, resulting from variations in brain development and circuitry.^1^ NDDs include intellectual developmental disorder (IDD), communication disorders, autism spectrum disorder (ASD), attention-deficit/hyperactivity disorder (ADHD), specific learning disorders, and developmental motor disorders.^1^ These conditions are highly heterogeneous but also often exhibit substantial overlap in their aetiology, biology, and phenotype.^2,3^

For individuals with NDDs and their families and caregivers, developmental impairments often result in significant barriers to activities of daily living, social participation, and quality of life, frequently lasting into adulthood.^4–6^ Among these challenges, communication difficulties are associated with poorer functional ability and quality of life and are considered a top priority for intervention by parents and caregivers.^7^ Speech-language pathologists (SLPs) are at the forefront of addressing these communication challenges, employing evidence-based strategies to diagnose and support overall communication abilities.^8^

The genetic underpinnings of many NDDs add a layer of complexity to diagnosis, prognosis, and intervention. Genetic NDDs, such as those involving 16p11.2 deletions/duplications, *ASXL3*, *CTNNB1*, *SCN2A*, *STXBP1*, *SYNGAP1*, and other genes, exhibit significant variability in their clinical presentations and developmental trajectories.^9–11^ While previous studies have investigated communication phenotype-genotype associations in NDDs, they were not explicitly focused on communication abilities, resulting in small sample sizes and limited phenotypic or genetic resolution. For example, one previous study relied on Social Communication Questionnaire (SCQ) Lifetime total and subdomain scores, such as the SCQ Social Interaction score, which could mask possible item-level variability within SCQ.^12^ Another study, using the September 2020 versions of the Simons Searchlight (Searchlight) and Simons Foundation Powering Autism Research (SPARK) registries, included only two communication milestones (ages at first used and combined words) and reported odds ratios without inferential statistics.^13^ Despite these limitations, both aforementioned studies provided compelling evidence for an association of communication abilities with genetically defined conditions.^12,13^ Studies of individual copy number variant (CNV)-based or monogenic NDD conditions, such as 16p11.2 deletion/duplication,^14^ *SCN2A*,^15^ and *STXBP1*,^16^ demonstrated that detailed analyses of communication abilities are feasible and can provide novel insights, but their single-condition focus precluded broader comparisons or scalable modelling of heterogeneity.

Supported by previous evidence, we hypothesised in this study that, among individuals with NDDs, communication abilities differ between individuals with a genetically caused NDD and those without known disease-associated mutations, as well as between specific genetic NDD conditions. We applied a harmonised analysis framework to fine-grained communication phenotypes in a large clinically curated NDD cohort (n = 79,518) to establish a foundation for integrating genetic testing results into speech-language pathology practice. We systematically analysed communication profiles at a subdomain- and measure-level resolution within a four-tiered comparison framework across genetic classes, individual genetic conditions, and age-stratified groups. Rather than relying on broad composite scores, we focused on lower-level communication indicators, including Vineland-3 expressive and receptive subdomains and selected milestone- and item-level measures from caregiver-report instruments, to capture fine-grained differences in verbal and nonverbal communication abilities that may remain elusive when employing exclusively scale-level scores (e.g., Vineland-3 Communication domain, SCQ total score). Using this approach, we analysed 10 key communication measures drawn from Vineland-3,^17^ an autism-related milestones questionnaire, SCQ,^18^ and the Social Responsiveness Scale, Second Edition (SRS-2)^19^ representing expressive, receptive, social, and nonverbal communication abilities, comprising 176,949 data points across the study cohorts. The findings expand and amend previous evidence^12,13^, demonstrate genetic condition-and age-specific communication profiles, and underscore the potential of precision SLP approaches to optimise interventions, address developmental delays, and improve outcomes.

## Methods

### Ethics

All participants (or their legal guardians) provided written informed consent to participate in the Searchlight^20^ and the SPARK^21^ studies, from which we drew the case and control cohorts for this study. The project was approved by the Simons Foundation (Project ID: 15099.1) and the institutional review board at UTHealth Houston (IRBs: HSC-MS-24-0469 and HSC-MS-23-1129).

### Communication outcomes

Within the Simons Foundation research projects, caregivers of participants completed standardised assessments covering multiple domains of communication. From these instruments, we selected 10 communication-related measures based on their (i) availability in both Searchlight and SPARK studies, (ii) direct relationship to core expressive, receptive, and social communication behaviours, and (iii) applicability across a wide developmental age range (Table 1). For clarity, throughout this manuscript, the term “verbal communication” refers to oral spoken communication, whereas nonverbal communication includes gesture-based and other non-oral communicative skills. From the Vineland-3 Parent/Caregiver Form,^17^ we extracted the communication domain subscale scores for Expressive and Receptive communication (as standardised v-scale scores with a mean of 15 and standard deviation of 3), rather than the higher-order Communication domain score, as these subscale scores capture oral communication and without dilution by written communication items. In contrast, for caregiver-report instruments such as the Searchlight/SPARK Milestones questionnaire, SCQ, and SRS-2, we selected specific items related to clearly defined communication behaviours, enabling finer-grained characterisation of communication profiles than total or composite scores. We extracted the age at which the child first used single words and first combined words into phrases from the Background History Questionnaires of the Searchlight and SPARK studies, which are informed by established developmental frameworks^22^ (Milestone-Q8 and -Q11). The milestone measurements were set to missing if: 1.) ages at which milestones were achieved were above the age at evaluation; 2.) age used words < 8 months;^23^ 3.) age combined words < 16 months.^23,24^ Individuals documented to combine words before first using words had Milestone-Q11 set to missing. From the SCQ Lifetime form,^18^ we extracted Q01 (key concept: Using phrases), Q02 (Having conversation), Q23 (Using gestures), Q24 (Nodding head), and Q25 (Shaking head). Finally, from SRS-2,^19^ we extracted Q12 (Expressing feelings) (Table 1). For all 10 measures, we removed individual-level scores flagged with questionable validity in the Searchlight and SPARK registries.

**Table 1.** Selected communication-related measures.

| Instrument | Item | Content | Key concept | Category | Type: Measurement |
| --- | --- | --- | --- | --- | --- |
| Vineland-3 Parent/Caregiver Form | Expressive subdomain | Assesses expressive language, including spoken and alternative language modalities (49 items) | Expressive | Verbal | Continuous: norm-referenced v-scale score |
|  | Receptive subdomain | Assesses receptive language, including understanding of spoken language and communicative signals (39 items) | Receptive | Comprehension-based* |  |
| Milestones | Q08 | Age used words | Age used words | Verbal | Time-to-event: age at milestone achievement |
|  | Q11 | Age combined words | Age combined words | Verbal |  |
| SCQ Lifetime form | Q01 | Is she/he now able to talk using short phrases or sentences? If no, skip to question 8. | Using phrases | Verbal | Binary: Yes/No |
|  | Q02 | Do you have a to and fro “conversation” with her/him that involves taking turns or building on what you have said? | Having conversation | Verbal |  |
|  | Q23 | Does she/he ever use gestures, other than pointing or pulling your hand, to let you know that she/he wants? | Using gestures | Nonverbal |  |
|  | Q24 | Does she/he nod her/his head to indicate yes? | Nodding head | Nonverbal |  |
|  | Q25 | Does she/he shake her/his head to indicate no? | Shaking head | Nonverbal |  |
| SRS-2 | Q12 | Is able to communicate his or her feelings to others | Expressing feelings | Social* | Ordinal: 1 = Not true; 2 = Sometimes true; 3 = Often true; 4 = Almost always true |

Shown are the ten caregiver-reported communication measures selected from Vineland-3, speech milestones, the Social Communication Questionnaire (SCQ), and the Social Responsiveness Scale, Second Edition (SRS-2). Measures were categorised according to their primary communication construct as verbal, nonverbal, comprehension-based, or social communication. Comprehension-based (Vineland-3 Receptive) and social communication (SRS-2 Q12) measures are marked with an asterisk (*) to indicate that they assess modality-agnostic communicative functions that may be expressed through verbal or nonverbal means.

### Study cohorts: discovery case-control cohort

The study included de-identified data from participants enrolled in the Searchlight^20^ and SPARK^21^ studies. Searchlight^20^ is a genetics-first research study that recruited participants with pathogenic or likely pathogenic (P/LP) variants in known NDD-associated genes or on recurrent CNVs, as curated by the Simons Foundation (Table 2).^25^ For our *Discovery* cohort, we selected individuals with the most prevalent genetic etiologies from the Searchlight registry v13.0: genetic conditions with more than 50 enrolled unique individuals with data available across the 10 communication-related measures chosen for this study, before quality control [Table 2, *Discovery: Genetic NDDs (prevalent forms)*]. This resulted in 15 distinct genetic conditions, including four recurrent CNVs^26,27^ and 11 monogenic conditions.^28–38^ As controls, we selected individuals with NDD from the SPARK study that used a “phenotype-first” approach to recruit participants with a diagnosis of autism spectrum disorder (ASD) made by healthcare professionals. We excluded all individuals whose ASD diagnosis was not confirmed and documented in the SPARK registry. We also excluded all individuals with reported genetic variants or CNVs identified through the clinical genetic testing within the SPARK study [Whole Exome Sequencing or CNV screening], using the September 15, 2025, data release, complemented by self-reported genetic diagnoses (Table 2, *Controls: Idiopathic ASD*). Because all control individuals had a confirmed or documented ASD diagnosis but no identified large-effect variants, the cohort is enriched with idiopathic, autism-associated NDD, and we refer to it as “idiopathic ASD controls” throughout the paper. However, only ∼50% SPARK participants received WES; thus, come controls may carry undetected genetic risk. A total of 335 unaffected siblings from the Searchlight registry were available to provide contextual reference values for communication behaviours and visualisations, but were used only in statistical testing for SRS-2, due to limited sample sizes for all other selected measures (Table 2).

**Table 2.** Study cohorts after quality control.

| Main analysis groups | Number of individuals | % male | % ASD | % Epilepsy or seizures | % Intellectual disability | Mean age walked [months] | % not walked by 7 years | Average household income [\$]‡ |
| --- | --- | --- | --- | --- | --- | --- | --- | --- |
| Discovery: Genetic NDDs (prevalent forms) | 1,521 | 54.1 | 38.1 | 42.2 | 40.8 | 22.6 | 3.52 | 93,306 |
| Confirmation-1: Genetic NDDs (rare forms) | 1,312 | 55.9 | 44.7 | 27.6 | 41.2 | 21.5 | 3.08 | 96,327 |
| Confirmation-2: Genetic ASD forms | 1,606 | 66.5 | 100 | 15.7* | 55.1 | 18.3 | 0.24 | 86,336 |
| Controls: Idiopathic ASD | 75,079 | 76.3 | 100 | 4.18* | 23.5 | 14.1 | 0.02 | 74,748 |
| Unaffected siblings† | 335 | 58.5 | 0 | 0 | 0.78 | 29.7 | 0 | - |
| Discovery cohort individuals grouped by genetic NDD condition | Number of individuals | % male | % ASD | % Epilepsy or seizures | % Intellectual disability | Mean age walked [months] | % not walked by 7 years | Average household income [\$]‡ |
| 16p11.2 deletion | 246 | 59.8 | 42.9 | 34.9 | 21.2 | 16.8 | 0 | 92,721 |
| PPP2R5D | 143 | 53.8 | 26.6 | 43.4 | 37.9 | 28.3 | 5.13 | 91,590 |
| CTNNA1 | 133 | 55.6 | 16.3 | 9.09 | 46.2 | 35.9 | 14.3 | 87,667 |
| SCN2A | 132 | 52.3 | 58.3 | 66.1 | 54.6 | 19.8 | 1.15 | 95,021 |
| 16p11.2 duplication | 126 | 57.1 | 44.3 | 26.4 | 33.0 | 15.8 | 0 | 75,743 |
| STXBPI | 126 | 48.4 | 34.3 | 80.0 | 45.5 | 34.6 | 19.4 | 98,057 |
| GRIN2B | 96 | 56.3 | 30.3 | 35.1 | 56.2 | 28.2 | 3.28 | 100,572 |
| SLC6A1 | 87 | 54.0 | 43.6 | 79.5 | 46.2 | 19.2 | 0 | 108,562 |
| CSNK2A1 | 83 | 44.6 | 19.7 | 37.0 | 39.4 | 27.3 | 1.41 | 93,676 |
| SYNGAP1 | 77 | 51.9 | 58.7 | 80.9 | 55.6 | 22.7 | 3.08 | 101,375 |
| MED13L | 71 | 52.1 | 31.6 | 28.6 | 66.7 | 25.5 | 0 | 106,851 |
| 1q21.1 duplication | 62 | 59.7 | 48.0 | 9.26 | 18.0 | 16.3 | 0 | 76,629 |
| 1q21.1 deletion | 53 | 52.8 | 30.4 | 17.4 | 13.0 | 14.2 | 0 | 73,642 |
| ASXL3 | 48 | 50.0 | 59.5 | 37.2 | 57.1 | 34.4 | 14.3 | 101,422 |
| SETBP1 | 38 | 50.0 | 28.0 | 22.2 | 60.0 | 20.5 | 0 | 108,297 |

Legend: ‡Household income was available as categorical income bands. For analysis, each individual's income was approximated by the midpoint of their reported band, and group-level values represent the mean of these midpoints. \*Percent from the total sample; the number of individuals without seizures or epilepsy was unknown in the SPARK registry. †Unaffected siblings were only used in statistical testing for SRS-2, due to limited sample sizes for all other selected measures.

### Study cohorts: confirmatory case cohorts

We then built two independent case cohorts and tested them against the same idiopathic ASD controls, thereby enabling partial validation of the results in the *Discovery* cohort (Table 2). We designed the “*Confirmation-1: Genetic NDDs (rare forms)*” cohort to test the *Discovery* cohort results, unbiased by the narrow selection of genetic NDD conditions in the *Discovery* cohort (n = 15), which may not be generalizable to other NDD conditions. The cohort consisted of all other genetic conditions in the Searchlight registry v13.0 that did not meet our inclusion criteria for the *Discovery* cohort (i.e., fewer than 50 unique individuals with data available before quality control; n = 109 monogenic conditions and n = 15 CNVs).

Previous analyses of the Searchlight cohort have shown that participants are strongly enriched for global developmental delay, IDD, and epilepsy, whereas autism diagnoses are present in only a subset of individuals (Table 2).^12,39^ Due to the lower rates of ASD in the *Discovery* and *Confirmation-1* cohorts, we expected both cohorts to be enriched for individuals with higher communicative intent compared to the idiopathic ASD control cohort,^40,41^ potentially driving differences in nonverbal communication (e.g., gestures). Therefore, we designed a second confirmation cohort (“*Confirmation-2: Genetic ASD forms*”) to determine whether discovery analysis results were generalizable to individuals with genetic ASD conditions, beyond those eligible for Searchlight.^25^ This cohort included individuals from SPARK with a confirmed clinical ASD diagnosis and pathogenic or likely pathogenic genetic variants affecting a broad spectrum of genes with strong clinical gene–phenotype validity for ASD, as detailed in “Study cohorts – quality control and age group definition” (n = 211 monogenic conditions; n = 28 distinct CNVs).

### Study cohorts: quality control and age group definition

All case cohorts (*Discovery*, *Confirmation-1*, *Confirmation-2*) were filtered to include only individuals carrying pathogenic (P) or likely pathogenic (LP) variants, in accordance with ACMG guidelines.^42^ We excluded individuals with variants of uncertain significance (VUS), considering the most recent variant evaluations in the “laboratory results” of the Searchlight v13.0 and SPARK September 15, 2025, data releases. For the *Discovery* and *Confirmation-1* cohorts, both consisting of Searchlight study participants,^20^ we included all P/LP variant carriers, as all affected genes or recurrent CNV regions were, by design of the Searchlight study, curated to be established genetic causes for NDD or ASD.^20^ For the *Confirmation-2* cohort, consisting of SPARK participants, we filtered genes based on the strength of evidence linking a gene to ASD, using the SFARI Gene database.^43,44^ We only included carriers of P/LP genetic variants affecting genes implicated in ASD with high confidence (SFARI gene categories 1 and 1S) or as strong candidates (SFARI gene categories 2 and 2S). CNVs were filtered to include only CNVs listed in SFARI Gene,^45^ or featured among eligible genetic conditions for Searchlight.^25^

Finally, all cohorts were filtered to include only individuals under 18 years of age at the time of evaluation to ensure consistency across measures and cohorts, following the design of previous Searchlight/SPARK studies.^13,46^ Individuals were also categorised into three age groups for cross-sectional analyses of developmental trajectories based on age at evaluation of each communication measure. The first age group, Preschool (0 to 5 years, 11 months), covered early development milestones.^24^ The other age groups were defined as Middle childhood (6 to 11 years, 11 months) and Adolescence (12 to 17 years, 11 months) based on standard developmental staging frameworks in paediatrics.^47^ After quality control, the *Discovery* cohort included 1,521 individuals, and the *Confirmation-1* and *Confirmation-2* cohorts included 1,312 and 1,591 individuals, respectively. The idiopathic ASD control cohort included 75,079 individuals (Tables 2 and 3).

### Statistical analysis

We first performed case-control comparisons to evaluate whether, among individuals with NDD, carriers of genetic causes (CNVs or monogenic mutations) differed in their communication abilities from those without known mutations. The three independent case cohorts (*Discovery*, *Confirmation-1*, and *Confirmation-2*) were each compared against a shared control group (*Controls: Idiopathic ASD*) (Table 2, Supplementary Table 1). Within the *Discovery* cohort, we also tested each of the 15 genetic conditions against the idiopathic ASD controls to identify communication profiles specific to each genetic condition. Each communication measure was analysed using the appropriate statistical model:

For Vineland-3, continuous outcomes (i.e., expressive and receptive v-scores) were analysed using Gamma generalised linear models with a log link adjusted for sex. Age at evaluation was not included as a covariate, as v-scores are standardised relative to age norms. For each comparison, we report the two-sided Wald *p*-value, the mean ratio (MR) as the effect size, and 95% Wald confidence intervals (CIs). The association results were visualised using the means of the continuous outcomes with t-based 95% CIs.

For the speech milestones (Milestone-Q8, Milestone-Q11), the data were modelled as time-to-event outcomes. We used Kaplan-Meier estimation to summarise the distribution of age at milestone attainment and calculated pointwise 95% CIs using Greenwood’s formula. The survival estimates and CIs were transformed into cumulative probabilities of having used or combined words by a given age. To evaluate group differences, we fitted Cox proportional hazards regression models, adjusted for sex. Age at evaluation was not included as a covariate because individuals were right-censored at their evaluation age if the milestone had not yet been achieved. For each testing group, we report the two-sided Wald *p*-value and the hazard ratio (HR) as the effect-size estimate from Cox models with 95% CIs. For visualisations, we used cumulative probabilities and 95% CIs for attaining the milestone at the maximum observed follow-up time.

For SCQ, dichotomous Yes/No outcomes of SCQ-Q01, -Q02, -Q23, -Q24, and -Q25 were analysed using binomial logistic regression with a logit link adjusted for sex and age at evaluation. For each comparison, we report the two-sided Wald *p*-value, the odds ratio (OR) as the effect size, and 95% Wald CIs for the ORs. For visualisations, we used group proportions (“Yes” rates) with Clopper-Pearson 95% CIs.

For the SRS-2 Q12 “Expressing feelings” (coded as: 1 = never/not true, 2 = sometimes, 3 = often, 4 = almost always), we modelled the ordinal outcome using proportional odds (cumulative logit) regression adjusted for sex and age at evaluation.^48,49^ For each comparison, we report the two-sided Wald *p*-value and the proportional odds ratio (POR) as the effect size with Wald 95% CIs. For visualisations, we used unadjusted means of the ordinal outcomes, with t-based 95% CIs computed on the numeric coding.

Finally, using the same statistical frameworks, we examined developmental trajectories through cross-sectional analyses of the previously defined age-at-evaluation groupings: Middle Childhood and Adolescence were each tested against the Preschool group. These age-group-stratified statistical tests, adjusted for sex, were performed separately for each genetic status category (*Discovery* cohort, the 15 genetic conditions in the *Discovery* cohort, both *Confirmation* cohorts, idiopathic ASD controls, and unaffected siblings for SRS-2 only). Age-group-stratified statistical tests were not performed for any testing group with fewer than five measurements. Multiple testing was addressed by Bonferroni correction, based on the number of case-control and age-group tests within each communication measure. The threshold for significant associations after Bonferroni correction for Vineland-3, speech milestones, and SCQ was set to α = 7.94×10^−4^, corresponding to 63 tests each. In addition to the Bonferroni-corrected significance threshold, we defined the threshold for suggestive associations as 1/number of tests (α = 0.016), a commonly used convention in genetic association studies to identify findings expected to occur by chance only once and therefore warranting further replication.^50,51^ For SRS-2, we performed 66 tests, resulting in α = 7.58×10^−4^ and α = 0.015 for significant and suggestive associations, respectively. All analyses were performed in RStudio version 2025.09.2 Build 418, using the stats (v4.3.2), survival (v3.5.7), and MASS (v7.3.60) R packages for statistical analyses.

### Data Sharing Statement

Approved researchers can obtain the Simons Searchlight (https://www.sfari.org/resource/simons-searchlight/) and the SPARK population dataset described in this study (https://www.sfari.org/resource/spark/) by applying at https://base.sfari.org.

## Results

### Overview of study participants and data points

This study analyzed 176,949 data points collected from 79,518 unique individuals with NDD, including 4,439 individuals with a CNV-based or monogenic NDD condition and 75,079 with idiopathic ASD (Fig. 1). Datapoints were gathered using Vineland-3 (n = 31,280), speech milestones (n = 49,055), SCQ (n = 76,820), and SRS-2 (n = 19,794) assessments (Supplementary Table 1). Across the matrix of the 10 communication measures evaluated across independent analysis groups (15 distinct genetic conditions of the *Discovery* cohort, and the two confirmation cohorts), 8 of the 10 measures showed strong and significant pairwise correlations (Spearman’s ρ > 0.8, *P* < 1.11×10^−3^). SCQ-Q23 (i.e., use of gestures) showed no significant correlation with any of the other measurements, and SRS-2-Q12 (i.e., ability to communicate feelings) was significantly correlated with only three measures (Supplementary Figure 1). Taken together, the correlation structure demonstrates that the selected communication measures encompass both overlapping and distinct facets of communication, with SCQ-Q23 capturing nonverbal and SRS-2-Q12 capturing social communication abilities that are largely independent of verbal communication in our cohort, thereby offering complementary information for delineating genotype-specific phenotypic communication profiles.

**Figure 1.**
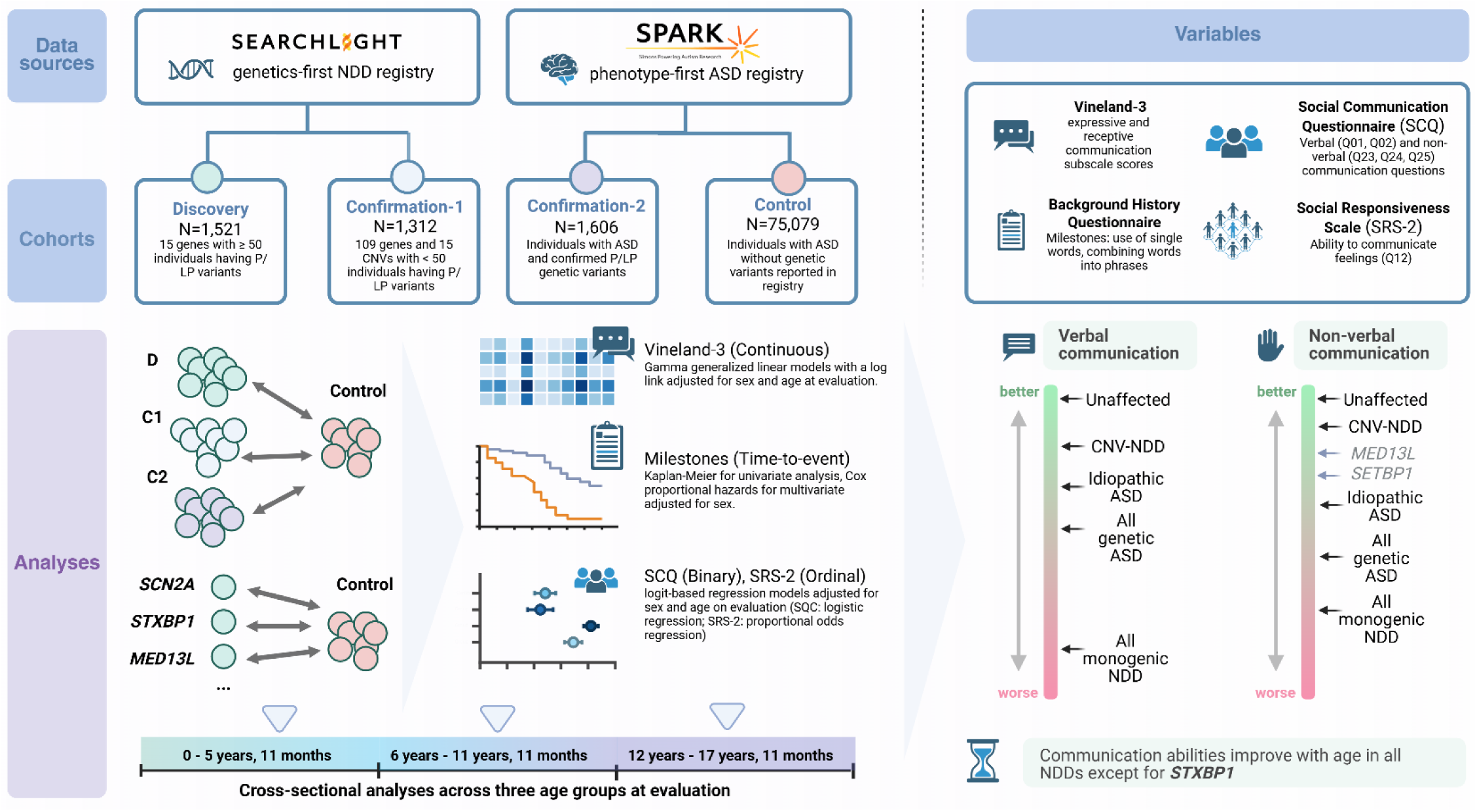
Study design. Communication outcomes from 4,439 individuals with neurodevelopmental disorders (NDDs) and pathogenic/likely pathogenic variants were organised into a discovery cohort and two independent confirmation cohorts, and tested against 75,079 idiopathic ASD controls (without known pathogenic variants). Ten verbal and nonverbal communication measures derived from Vineland-3, speech milestones, SCQ, and SRS- 2 were evaluated using outcome-appropriate statistical models. Analyses compared genetic classes, individual genetic conditions, and age-stratified groups to identify genotype-specific communication profiles and patterns of change with age.

### Genetic NDDs are associated with lower verbal and comprehension-based but higher nonverbal and social communication

Individuals with the most prevalent CNV-based or monogenic NDD conditions in the Searchlight registry (n = 15, *Discovery* cohort, Supplementary Table 1) showed, at the group level, significantly lower abilities on all five measures of verbal communication compared to idiopathic ASD controls, as well as the measure of comprehension-based communication. On average, their Vineland-3 v-scaled scores were 22% lower for expressive (*P*_Gamma-GLM_ = 4.53×10^−115^) and 17% lower for receptive (*P*_Gamma-GLM_ = 7.14×10^−58^) communication abilities. Early speech development was also markedly delayed, with 45% lower likelihood of producing first words (Milestone-Q8: *P*_Cox-PH_ = 8.17×10^−70^) and 44% lower likelihood of combining words into phrases (Milestone-Q11: *P*_Cox-PH_ = 4.40×10^−52^) at any given age. In parallel, the odds of exhibiting key verbal communication behaviors were significantly reduced, with 59% lower odds of using phrases (SCQ-Q01: *P*_binomial-GLM_ = 1.15×10^−39^) and 20% lower odds of engaging in back-and-forth conversation (SCQ-Q02: *P*_binomial-GLM_ = 6.44×10^−4^) (Fig. 2, Supplementary Figures 3-8, Supplementary Table 2). Interestingly, at the group level, the same individuals had higher scores on two measures of nonverbal communication and on the measure of social communication. They had significant increases in the odds of using gestures – 26% higher (SCQ-Q23: *P*_binomial- GLM_ = 1.12×10^−4^) and of expressing their own feelings appropriately – 29% higher (SRS-2-Q12: *P*_ordinal logistic regression_ = 1.93×10^−8^). A similar, though more modest, suggestive increase was observed in the odds of nodding “yes” (SCQ-Q24: *P*_binomial-GLM_ = 2.47×10^−3^) (Fig. 2, Supplementary Figures 9-12, Supplementary Table 2).

**Figure 2.**
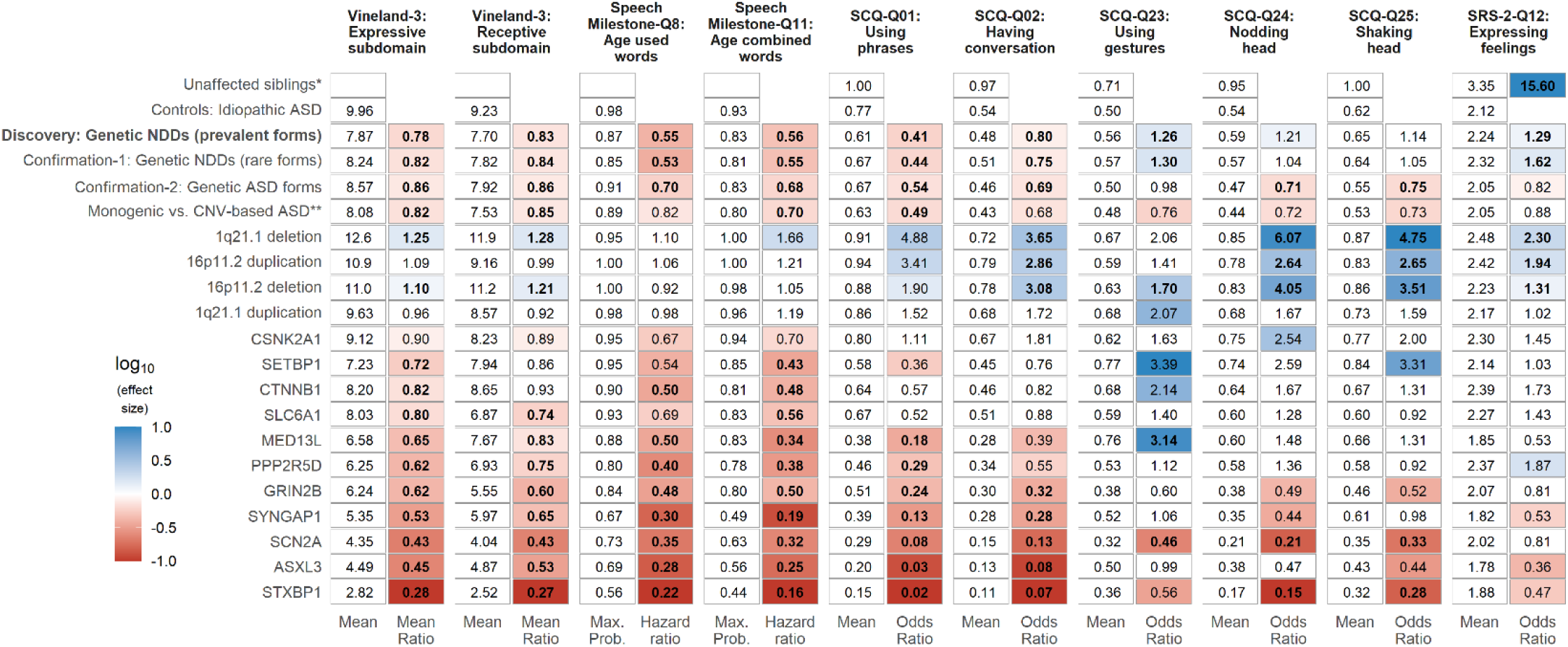
Group-level differences in communication measures across genetic NDD conditions. Shown are group-level means and corresponding effect sizes for 10 communication-related measures across three cohorts (Discovery, Confirmation-1, Confirmation-2) and 15 genetic NDD conditions, each compared to the idiopathic ASD control group. The effect sizes for significant and suggestive association signals are highlighted in red (indicating lower communication outcomes in each case cohort) and blue (indicating higher communication outcomes). Bolded values indicate statistically significant associations after multiple testing correction; non-bold colored values reflect suggestive associations. Legend: Max Prob.: maximum cumulative probability of milestone attainment at the latest observed follow-up; SCQ: Social Communication Questionnaire; SRS-2: Social Responsiveness Scale, Second Edition; *Outcome measures for unaffected siblings were only available for SRS- 2 in a sufficient size for statistical testing, where they were tested against idiopathic ASD controls for consistency. **Monogenic ASD forms tested against CNV-based ASD forms, both subsets of the Confirmation-2 cohort.

These core findings were partially replicated in two independent case cohorts, each drawn from distinct subsets of the Searchlight (*Confirmation-1*: rare genetic NDDs) and SPARK datasets (*Confirmation-2*: genetic ASD forms) (Tables 2 and 3). In both confirmation analyses, individuals with genetic NDD or ASD conditions again demonstrated significantly lower communication scores in verbal and comprehension-based communication measures (i.e., Vineland-3, speech milestones, SCQ-Q01, and SCQ-Q02) compared to the same idiopathic ASD controls (Fig. 2). However, we consider these analyses as only partial confirmations, as all analyses used the same idiopathic ASD control cohort, which limits their independence for replication. For the nonverbal and social communication measures, results were mixed across the two confirmation cohorts. The *Confirmation-1* cohort closely mirrored the *Discovery* cohort, replicating the significantly higher scores in SCQ-Q23 and SRS-2-Q12 (>30% higher odds, *P* < 1.58×10^−4^). In contrast, the Confirmation-2 cohort, comprising exclusively individuals with ASD (compared with 38% and 45% ASD in the Discovery and Confirmation-1 cohorts, respectively; Table 2), showed an opposite pattern on nonverbal communication measures and did not replicate the previously observed association for the social communication measure. Specifically, the cohort showed significantly lower odds of nodding “yes” – 29% lower (SCQ-Q24: *P*_binomial-GLM_ = 9.76×10^−11^) and of shaking head for “no” – 25% lower (SCQ-Q25: *P*_binomial-GLM_ = 2.86×10^−8^), indicating that the higher odds for using gestures (SCQ-Q23) and expressing feelings (SRS-2-Q12) observed in the *Discovery* cohort were not generalizable to individuals with genetic ASD conditions (Fig. 2, Supplementary Figures 10-11, Supplementary Table 2).

### Genetic NDDs have heterogeneous communication profiles

We then tested each of the 15 most prevalent CNV/based or monogenetic NDD conditions represented in the *Discovery* cohort against the control group (idiopathic ASD) to identify whether communication profiles are specific to each genetic condition. Results revealed striking genotype-specific heterogeneity in communication outcomes, with some genetic NDD groups showing substantial deviations in their mean communication scores.

Individuals carrying recurrent NDD-associated CNVs achieved the highest communication scores across all communication measures, often significantly exceeding those of idiopathic ASD controls. Across the 16p11.2 deletion, 16p11.2 duplication, and 1q21.1 deletion CNV groups, we observed higher scores in nine out of the 10 communication-related measures, with differences compared to idiopathic ASD controls reaching statistical significance in at least four communication measures for each CNV (1q21.1 deletion: six measures with 1.25 to 6.07-fold higher mean scores or odds; 16p11.2 duplication: four measures with 1.94 to 2.65-fold higher odds; 16p11.2 deletion: seven measures with 1.10 to 4.05-fold higher mean scores or odds) (Fig. 2, Supplementary Figures 3-12, Supplementary Table 2).

In contrast, all 11 monogenic NDD groups showed lower verbal and comprehension-based communication scores, each compared to idiopathic ASD controls, with differences in communication profiles revealed by the nonverbal and social communication measures. Five of these monogenic conditions displayed mixed communication profiles with lower scores in three to four of the six measures representing verbal and comprehension-based communication abilities but higher scores in at least one of the nonverbal or social communication measures compared to idiopathic ASD controls (three signals from SCQ-Q23 and one each from SCQ-Q24, SCQ-Q25, and SRS-2-Q12) (Fig. 2, Supplementary Table 2). The results for SCQ-Q23 are consistent with the absence of significant correlations with other measures, demonstrating that SCQ-Q23 can identify fine-grained differences that are missed by composite scores that include gestures (i.e., Vineland-3 expressive). The *SETBP1* and *MED13L* groups showed the two overall highest SCQ-Q23 mean scores, outperforming the odds of using gestures in the idiopathic ASD control group by 3.39-fold (*P*_binomial-GLM_ = 4.46×10^−3^) and 3.14-fold (*P*_binomial-GLM_ = 5.37×10^−4^), respectively (Supplementary Figure 9). Also, their ability to use gestures (mean percentages of “yes” answers in SCQ-Q23: 0.76-0.77) was on par with that of the 38 available unaffected siblings (mean percentage 0.71) (Supplementary Table 2). One gene group (*SLC6A1*) showed lower abilities on verbal communication measures and on the comprehension-based communication measure, whereas nonverbal communication abilities did not differ statistically from those of controls. The remaining five monogenic conditions (*GRIN2B*, *SYNGAP1*, *SCN2A*, *ASXL3*, and *STXBP1*) showed broadly reduced communication abilities across most communication measures, with *SCN2A, ASXL3*, and *STXBP1* emerging as the most severely affected groups - in line with the global developmental delays and neurological complications reported for these syndromes^16,52–54^ (*SCN2A*: 9/10 measures with significantly lower scores; *ASXL3*: 7/10 significant and 1/10 suggestive; *STXBP1*: 8/10 significant and 2/10 suggestive; Fig. 2).

### Monogenic NDDs exhibit lower communication abilities than CNV-based NDDs across all measures

To further contextualize and confirm genotype-specific differences in communication profiles observed in the *Discovery* cohort, we statistically tested communication outcomes of monogenic NDD against CNV-based NDD conditions in the *Confirmation-2* cohort, which included a broader representation of genetic conditions than the *Discovery* cohort. The pattern was clear and consistent: children with monogenic NDDs showed lower communication abilities across 9 of the 10 measures examined compared to CNV carriers. Significant differences were observed in four measures: monogenic NDDs had 18% lower expressive (*P*_Gamma-GLM_ = 1.19×10^−6^) and 15% lower receptive (*P*_Gamma-GLM_ = 7.36×10^−5^) mean Vineland-3 v-scaled scores, 30% lower likelihood of combining words into phrases (Milestone-Q11: *P*_Cox-PH_ = 1.38×10^−6^) at any given age, and had 51% lower odds of using phrases (SCQ-Q01: *P*_binomial-GLM_ = 4.42×10^−8^). Five other measures (Milestone-Q8 and SCQ-Q02/Q23/Q24/Q25) showed suggestive differences (*P* < 0.011) (Fig. 2, Supplementary Figures 3-12, Supplementary Table 2). Only one measure, SRS-2-Q12, did not differ statistically between groups. This group-level analysis complements the genetic condition-specific analyses in the *Discovery* cohort, reinforcing the observation that monogenic NDDs are associated with broader and more severe communication impairments than CNV-based NDDs, both in absolute terms and relative to individuals with idiopathic ASD.

### Age-stratified developmental trajectories: general improvement except for STXBP1

We next performed cross-sectional analyses across three age groups at evaluation to determine whether developmental trajectories differ by genetic status. Among individuals with idiopathic ASD, we observed a consistent pattern of improvement in communication abilities with increasing age at evaluation. Specifically, the idiopathic ASD controls showed significant increases in abilities from the “Preschool” (0 to 5 years, 11 months) to the “Middle Childhood” (6 to 11 years, 11 months) group in all measures. These increased abilities were maintained or further improved in the Adolescence group (12 to 17 years, 11 months) across 9 of the 10 communication measures examined, with the exception of SCQ-Q23 (Fig. 3; Supplementary Table 3). We then restricted our further interpretation to monotonic increases or decreases in mean scores across age groups, because non-monotonic patterns are more likely to reflect sampling variability than genuine late-onset developmental regression or early plateau effects. The strongest improvement of the idiopathic ASD controls was observed in SCQ-Q01, with 6.28-fold higher odds of using phrases in “Middle Childhood” and 10.21-fold higher odds in “Adolescence”. Notably, this monotonic increase was also observed for the SRS-2 Q12 mean scores, which, in typically developing individuals, usually decrease with increasing chronological age,^55^ as shown by the unaffected sibling group (mean ordinal outcomes of 3.55, 3.39, and 3.09 in “Preschool”, “Middle Childhood”, and “Adolescence” [*P*_ordinal logistic regression_ = 7.57×10^−4^], respectively) (Fig. 3 and Supplementary Figure 20), possibly reflecting increased self-consciousness in emotional expression among typical teenagers. All case cohorts mirrored the results in the idiopathic ASD controls: 1.) *Discovery* cohort (most prevalent genetic NDD conditions in Searchlight): 4 out of 10 communication-related measures with significant and monotonic increases from “Preschool” to “Adolescence”; 2.) *Confirmation-1* cohort (rare genetic NDD conditions in Searchlight): 4/10 communication-related measures with monotonic increases, supported by at least one suggestive difference, and 3.) *Confirmation-2* cohort (genetic ASD forms from SPARK): 8/10 communication-related measures with monotonic increases (Fig. 3).

**Figure 3.**
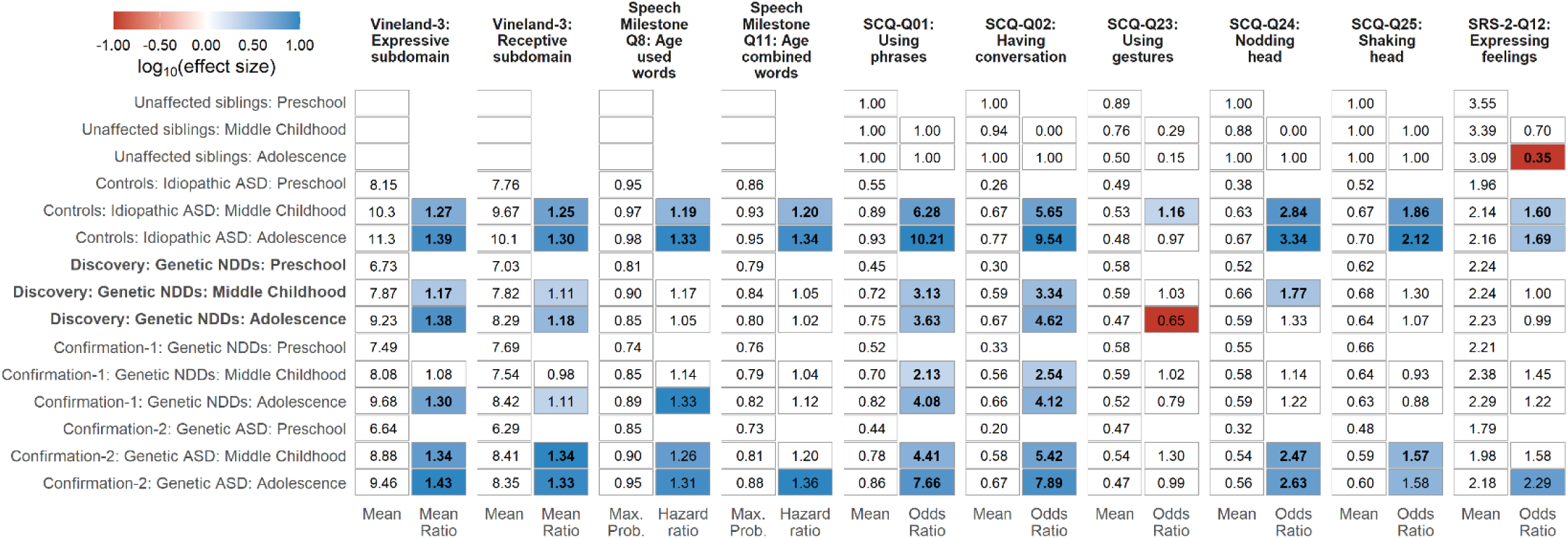
Age-stratified developmental trajectories in the Discovery, Confirmation-1, and Confirmation-2 cohorts. Shown are the cross-sectional age-stratified analyses of age at evaluation groups: “Middle Childhood” (6 to 11 years, 11 months) and “Adolescence” (12 to 17 years, 11 months), each tested against “Preschool” (0 to 5 years, 11 months). The effect sizes for significant and suggestive association signals are highlighted in red (indicating lower communication outcomes than the “Preschool” group) and blue (indicating higher communication outcomes). Bolded values indicate statistically significant differences between communication outcomes after multiple testing correction; non-bold colored values reflect suggestive differences. Legend: Max Prob.: maximum cumulative probability of milestone attainment at the latest observed follow-up; SCQ: Social Communication Questionnaire; SRS-2: Social Responsiveness Scale, Second Edition.

At the level of specific genetic NDD conditions (in the *Discovery* cohort), we observed at least one measure with at least one suggestive difference between age at evaluation groups, supporting a monotonic trend of increased abilities in 5 out of 15 genetic conditions: 16p11.2 duplication, 16p11.2 deletion, *CSNK2A1*, *SYNGAP1*, and *SCN2A*. These improvements were all captured, among other measures, by SCQ-Q01 (6.85 to 8.93-fold higher odds of using phrases in “Adolescence”, *P* < 0.014) and SCQ-Q02 (4.57 to 29.6-fold higher odds of using phrases in “Adolescence”, *P* < 0.012) (Supplementary Table 3, Supplementary Figures 2, 13-19).

The *STXBP1* group was the sole outlier that failed to follow the improvement trajectory observed in the *Discovery*, *Confirmation-1*, and *Confirmation-2* cohorts, as well as in several single conditions. Individuals with *STXBP1* mutations showed a monotonic decline in norm-referenced communication scores on the Vineland-3. Their expressive v-scale scores declined from 3.63 in “Preschool” to 2.20 in “Middle Childhood” (*P*_Gamma-GLM_ = 5.89×10^−3^) and 2.04 in “Adolescence” (*P*_Gamma-GLM_ = 2.57×10^−3^) (Supplementary Figures 2 and 13). Similarly, receptive mean v-scale scores declined significantly from 3.50 in “Preschool” to 1.84 in “Middle Childhood” (*P*_Gamma-GLM_ = 5.90×10^−4^) and 1.46 in “Adolescence” (*P*_Gamma-GLM_ = 1.29×10^−4^) (Supplementary Figures 2 and 14, Supplementary Table 3), indicating that many *STXBP1* mutation carriers may experience a plateau in communication skill development relative to age expectations, with the possibility that some individuals fail to acquire additional skills or, in some cases, lose previously acquired abilities. Consistent with this interpretation, Vineland-3 expressive and receptive raw scores for the *STXBP1* group were not statistically different between age groups (P > 0.04), supporting a pattern of developmental plateau; this contrasts with the previously reported pattern in *SCN2A* variant carriers, who appear to have increasing Vineland-3 raw scores with increasing age but more slowly than peers, leading to declining norm-referenced v-scaled scores.^56^

## Discussion

This study demonstrates considerable variability in communication abilities among individuals with genetic NDDs, underscoring the need to integrate genetic insights into speech-language pathology (SLP) practices. Specifically, we found that individuals with genetic NDD conditions performed significantly worse than those with idiopathic ASD across all measures of verbal and comprehension-based communication abilities. In contrast, they performed better on three of four measures assessing nonverbal and social communication abilities. Verbal and comprehension-based communication findings were replicated across both confirmation cohorts, whereas nonverbal and social communication findings were confirmed in only one cohort, indicating that this result is not generalizable to individuals with ASD.

By analysing each genetic NDD condition against the idiopathic ASD controls, we show that communication abilities differ systematically across genetic classes (monogenic vs. CNV-based NDDs) and across individual genetic diagnoses. CNV-based NDDs generally performed better than idiopathic ASD controls across measures of verbal, comprehension-based, and nonverbal communication, whereas all monogenic NDD conditions showed marked verbal and comprehension-based communication impairments. These observed differences between monogenic and CNV-based NDDs are in line with previous evidence from verbal communication data (i.e., language milestones^13^ and SCQ^12^). Within the monogenic groups, however, several conditions showed relative strengths in specific nonverbal skills for which, to our knowledge, no prior statistical evidence has been published. For example, individuals with *SETBP1* and *MED13L* variants demonstrated statistically higher gesture use (SCQ-Q23) than those with idiopathic ASD. Their ability to use gestures was comparable to that of unaffected siblings; however, this observation requires replication in a larger unaffected cohort. Interestingly, non-statistical evidence from a study reporting “intact non-verbal gestures” in individuals with *SETBP1*^57^ mutations and indirect evidence of augmentative and alternative communication (AAC) use in 7/17 children with *MED13L* mutations,^58^ could be considered as supportive of our results. The existence of genetic NDD conditions with impaired verbal but relatively high nonverbal communication abilities likely explains why a previous study failed^12^ to identify statistical differences between genetic NDDs and idiopathic ASD when using the SCQ total and the Social subdomain score, which both combine items such as having conversations (Q02) and gesture use (Q23).^59^

Encouragingly, cross-sectional age-stratified analyses revealed signs of communication gains with increasing age. At the group level, several communication abilities showed monotonic increases in five of the 15 conditions examined in depth (16p11.2 duplication, 16p11.2 deletion, *CSNK2A1*, *SYNGAP1*, and *SCN2A*). These improvements were particularly reflected in SCQ Q01 (“Using phrases”) and Q02 (“Having conversation”), indicating growth in expressive and reciprocal communication abilities. These patterns highlight continued developmental potential and support sustained SLP intervention across development. Among all investigated NDD conditions, *STXBP1* stands out by showing declining norm-referenced communication abilities on Vineland-3 with increasing age, confirming previous evidence suggesting a slowly progressive course of the disease in at least some patients with *STXBP1* mutations, with 26% of individuals with *STXBP1* mutations displaying loss of skills, mainly in the verbal and motor domains, beyond early childhood.^52^ Dedicated longitudinal studies will be essential for characterising communication trajectories in *STXBP1* and informing treatment.

Genetic diagnoses inform SLP care in two complementary ways: they guide intervention selection by aligning therapy with syndrome-specific communication strengths and weaknesses, and they provide prognostic value by revealing condition-specific developmental trajectories. For example, the *STXBP1* group showed age-related decline in normative communication scores, supporting early and sustained intervention. In contrast, several other genetic conditions demonstrate measurable improvement over time, indicating potential for gradual developmental gains.

### Incorporating Genetic Profiles into SLP Interventions

The findings underscore the importance of genetic information in informing SLP interventions.^60^ For example, SLPs working with individuals affected by CNV-based NDDs can build on the relatively strong expressive, receptive, and social-communication abilities observed in this study by targeting grammatical structures and conversational fluency in both structured and natural contexts. Such approaches may further strengthen social-pragmatic language skills, which have been reported as delayed in most verbal individuals with 16p11.2 CNVs.^14^ Conversely, for individuals with *SCN2A*, *ASXL3*, and *STXBP1* who exhibited the most significant impairments among the studied communication measurements, early adoption of assistive technology, such as AAC systems, may be critical to provide a foundation for communicative interaction while fostering social participation/engagement and promoting inclusive education.^61^ Therapies focused on basic expressive and receptive language skills, supported by caregiver-mediated interventions, could further address these foundational needs.^62,63^ For genetic NDD conditions with impaired verbal communication and relative strengths in nonverbal communication, such as *SETBP1* and *MED13L*, incorporating gestures into SLP intervention could facilitate communication and enhance social interaction,^64^, as evidence shows that caregivers are highly sensitive to both typical and unconventional nonverbal cues even when verbal communication is markedly limited.^65,66^

Genotype–phenotype associations are highly relevant to SLP practice, as several genetic NDD conditions showed distinct developmental trajectories, underscoring the need to tailor interventions to condition-specific developmental profiles. For example, the *SCN2A* group showed profound communication impairments across measures, consistent with previous evidence,^54^ but also modest age-related gains in conversational skills, which have not been described before, unlike the trajectory observed in the *STXBP1* group.

### Challenges to Implementing Precision SLP Approaches

Despite the promise of integrating genetic insights into SLP practices, several challenges must be addressed:

1. Genetic Literacy: Many SLPs lack formal training in genetics,^67^ making it challenging for them to incorporate genetic findings into clinical care. The diversity of SLP training pathways amplifies this challenge, as clinicians vary widely in their exposure to complex medical, neurodevelopmental, and genetic conditions. In this context, expanded professional development opportunities and potentially advanced training tracks or credentialed specialisations focused on genetic and neurodevelopmental disorders could better equip SLPs to deliver genetics-informed care.^60^ Interdisciplinary collaboration with specialists, such as genetic counsellors, will also support SLPs in understanding and utilising genetic information effectively.
2. Variability in Outcomes: Although group-level trends were identified in this study, substantial interindividual variability persists within genetic NDDs, reflecting the known variable expressivity of many of the analysed genes.^68,69^ Communication outcomes and therapy response may also be shaped by co-occurring cognitive and neurobehavioral factors, such as attention, executive functioning, motor impairments, or fatigue, underscoring the need for individualised application of group-level insights.
3. Access to Resources: Many families face barriers to accessing specialised care due to geographic, financial, and systemic constraints. Scalable solutions such as telehealth services, community-based programs, and equitable access to genetic testing and therapeutic services are essential to overcoming these challenges. Expanding access will require integrating genetic testing into routine SLP diagnostic workups for complex NDDs, coupled with advocacy for insurance coverage and reimbursement pathways that reduce financial barriers for families.

### Future Directions and Recommendations

Using our gene-level insight, future natural history and interventional studies should be stratified by genotype and specific clinical presentation, as exemplified by a previous study that investigated developmental outcomes in individuals with *STXBP1*,^70^ thereby informing the development of standardised approaches for incorporating genetic insights into SLP practice. Strengthening collaboration among SLPs, genetic counsellors, neurologists, paediatricians, and other pediatric specialists (e.g., neuropsychologists, psychologists, behaviour therapists) will improve the integration of genetic findings into clinical care, potentially including telehealth services to ensure equitable access across diverse populations.^71,72^

### Limitations of this study

Individuals with NDD in this study are self-referred participants in the Simons Foundation studies and may not represent the full range of possible communication-phenotype impairments. Individuals in the *Discovery* cohort were recruited using a genotype-first approach, which may have biased the cohort toward delays in early milestones that would have prompted genetic testing. We cannot report complete replications of our results, as all three case cohorts were tested against the same control cohort. Future studies would also benefit from control cohorts with a broader representation of NDD conditions, beyond the SPARK cohort, which is enriched with individuals with ASD. Previous evidence suggests that genotypes do not fully explain the phenotypic variability in communication abilities within a genetic group.^12,73^ Unmodeled factors such as exposure to SLP therapies, seizure burden, comorbid conditions, and genetic variant type may influence communication outcomes and warrant investigation in future studies. Finally, the term “developmental trajectories” should be interpreted with caution, as it refers here to cross-sectional age-stratified comparisons rather than within-individual longitudinal change. Future longitudinal studies will be essential for accurately characterising communication trajectories and informing treatment.

## Conclusion

By analysing >75,000 individuals with NDD, this study is the first large-scale investigation to demonstrate systematic differences in communication profiles across multiple genetic NDD conditions, underscoring the feasibility of a genetics-informed approach in SLP. Our results indicate that several conditions exhibit measurable gains with age, highlighting the need for longitudinal, developmentally informed care. By integrating genetics-first phenotyping with standardised communication inventories at scale, our study provides the foundation for future research that will ultimately lead to genetics-informed SLP interventions. Importantly, our findings support the use of genetically informed communication outcome measures in clinical trials, facilitating more precise assessment of treatment effects across distinct genetic NDD conditions. Looking ahead, we anticipate that incorporating genetic profiles into clinical decision-making and therapeutic planning in SLP will become essential to improving treatment precision and outcomes for individuals with NDD.

## Acknowledgements

We are grateful to all of the families participating in SPARK and Simons Searchlight, the SPARK clinical sites and staff, and the participating Simons Searchlight sites and consortium (formerly the Simons VIP Consortium).

## Contributors

CH and CL designed the study. CH, CL, and TB analysed the data. CH, CL, DL, and TB accessed and verified the underlying data, interpreted the analyses, and were responsible for the decision to submit the manuscript. DL provided the computational infrastructure. DL supervised the study. CH, CL, and DL wrote the manuscript. All authors interpreted the data, read and revised the manuscript, and approved the final version of the manuscript.

