## Supplementary Table 1 for "Genotype-specific communication profiles in 79,518 individuals with neurodevelopmental disorders"

**Corresponding author:**

Costin Leu, PhD

**1. Supplementary Tables**

1.1. Supplementary Table 1: Sample sizes and availability of communication-related measures across analysis cohorts and genetic conditions

**2. Supplementary Figures**

2.1. Supplementary Figure 1: Spearman's pairwise correlation analysis of the 10 communication-related measures across independent testing groups
2.2. Supplementary Figure 2: Age-stratified developmental trajectories across all testing groups
2.3. Supplementary Figure 3: Association analyses for Vineland-3 Expressive subdomain
2.4. Supplementary Figure 4: Association analyses for Vineland-3 Receptive subdomain
2.5. Supplementary Figure 5: Cumulative attainment of speech milestone Q8: "Age Used Words" across testing groups
2.6. Supplementary Figure 6: Cumulative attainment of speech milestone Q11: "Age Combined Words" across testing groups
2.7. Supplementary Figure 7: Association analyses for SCQ Q01 "Using phrases"
2.8. Supplementary Figure 8: Association analyses for SCQ Q02 "Having conversation"
2.9. Supplementary Figure 9: Association analyses for SCQ Q23 "Using gestures"
2.10. Supplementary Figure 10: Association analyses for SCQ Q24 "Nodding head"
2.11. Supplementary Figure 11: Association analyses for SCQ Q25 "Shaking head"
2.12. Supplementary Figure 12: Association analyses for SRS-2 Q12 "Expressing feelings"
2.13. Supplementary Figure 13: Age-stratified developmental trajectories for Vineland-3 Expressive subdomain
2.14. Supplementary Figure 14: Age-stratified developmental trajectories for Vineland-3 Receptive subdomain
2.15. Supplementary Figure 15: Age-stratified developmental trajectories for SCQ Q01 "Using phrases"
2.16. Supplementary Figure 16: Age-stratified developmental trajectories for SCQ Q02 "Having conversation"
2.17. Supplementary Figure 17: Age-stratified developmental trajectories for SCQ Q23 "Using gestures"
2.18. Supplementary Figure 18: Age-stratified developmental trajectories for SCQ Q24 "Nodding head"
2.19. Supplementary Figure 19: Age-stratified developmental trajectories for SCQ Q25 "Shaking head"
2.20. Supplementary Figure 20: Age-stratified developmental trajectories for SRS-2 Q12 "Expressing feelings"

### 1. Supplementary Tables

#### Supplementary Table 1: Sample sizes and availability of communication-related measures across analysis cohorts and genetic conditions

| Main analysis groups | Unique individuals | Vineland-3 | Speech milestones | SCQ | SRS-2 | TOTAL measurements |
|---|---|---|---|---|---|---|
| Discovery: Genetic NDDs (prevalent forms) | 1,521 | 1,748 | 1,295 | 1,155 | 1,969 | 6,167 |
| Confirmation-1: Genetic NDDs (rare forms) | 1,312 | 1,404 | 1,191 | 868 | 773 | 4,236 |
| Confirmation-2: Genetic ASD forms | 1,606 | 863 | 1,231 | 1,572 | 550 | 4,216 |
| Controls: Idiopathic ASD | 75,079 | 27,265 | 45,338 | 73,225 | 16,502 | 162,330 |
| Unaffected siblings | 335 | 0 | 3 | 38 | 338 | 379 |
| TOTAL individuals with NDD | 79,518 | 31,280 | 49,055 | 76,820 | 19,794 | 176,949 |
| **Discovery cohort individuals grouped by genetic NDD condition** | **Unique individuals** | **Vineland-3** | **Speech milestones** | **SCQ** | **SRS-2** | **TOTAL measurements** |
| 16p11.2 deletion | 246 | 277 | 213 | 210 | 655 | 1,355 |
| PPP2R5D | 143 | 211 | 118 | 104 | 103 | 536 |
| CTNNB1 | 133 | 128 | 124 | 76 | 54 | 382 |
| SCN2A | 132 | 102 | 97 | 97 | 109 | 405 |
| 16p11.2 duplication | 126 | 135 | 108 | 110 | 316 | 669 |
| STXBP1 | 126 | 128 | 101 | 88 | 49 | 366 |
| GRIN2B | 96 | 119 | 78 | 69 | 82 | 348 |
| SLC6A1 | 87 | 102 | 77 | 63 | 71 | 313 |
| CSNK2A1 | 83 | 107 | 77 | 61 | 74 | 319 |
| SYNGAP1 | 77 | 69 | 68 | 54 | 51 | 242 |
| MED13L | 71 | 92 | 66 | 50 | 20 | 228 |
| 1q21.1 duplication | 62 | 86 | 52 | 56 | 156 | 350 |
| 1q21.1 deletion | 53 | 89 | 46 | 46 | 153 | 334 |
| ASXL3 | 48 | 55 | 33 | 40 | 40 | 168 |
| SETBP1 | 38 | 48 | 37 | 31 | 36 | 152 |

Shown are available communication-related measures across analysis cohorts from the Vineland Adaptive Behavior Scales Third Edition (Vineland-3 Expressive and Receptive subdomains), speech milestones from the Background History Questionnaires of the Searchlight and SPARK studies (Q8 and Q11), Social Communication Questionnaire (SCQ Q01, Q02, Q23, Q24, and Q25), and the Social Responsiveness Scale, Second Edition (SRS-2 Q12).

#### 2. Supplementary Figures

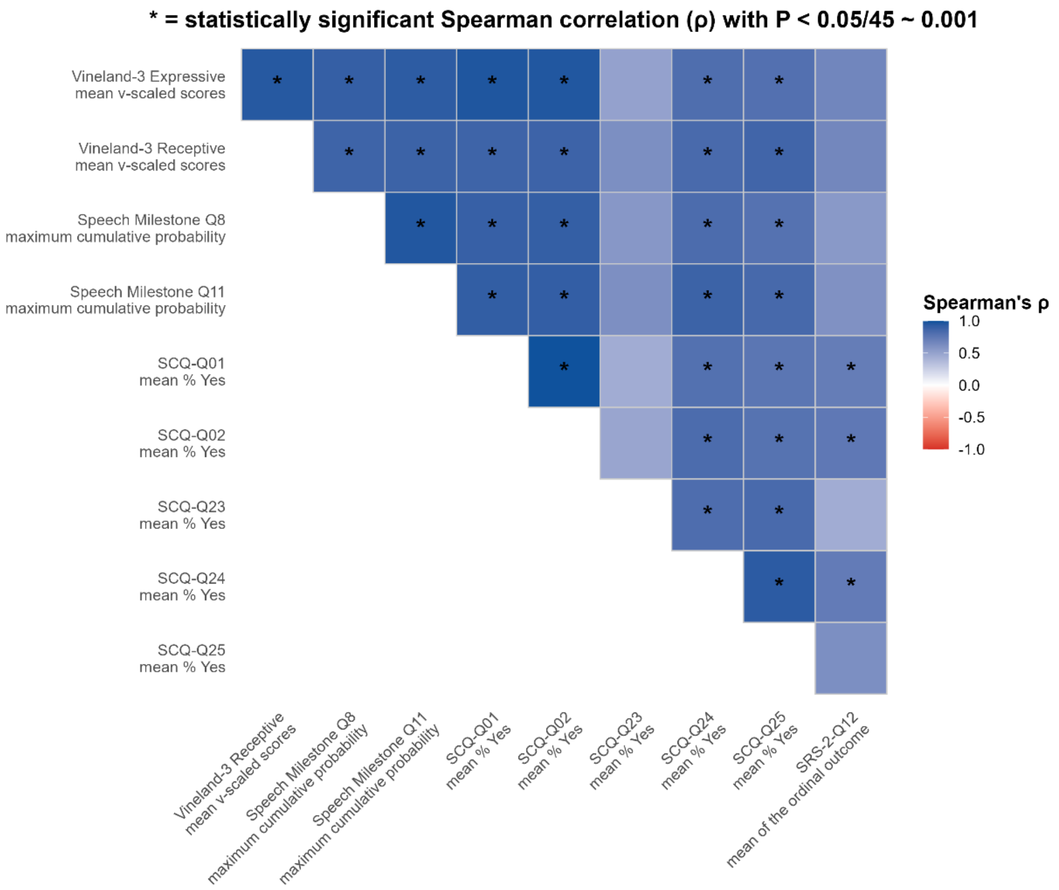


**Supplementary Figure 1: Spearman's pairwise correlation analysis of the 10 communication-related measures across independent testing groups**

Positive correlation coefficients (Spearman's ρ) are coloured in blue. Asterisks indicate correlations significant after Bonferroni multiple-testing correction (45 tests).

| Group | Vineland-3: Expressive subdomain: Mean | Vineland-3: Expressive subdomain: Mean Ratio | Vineland-3: Receptive subdomain: Mean | Vineland-3: Receptive subdomain: Mean Ratio | Speech Milestone Q8: Age used words: Max. Prob. | Speech Milestone Q8: Age used words: Hazard ratio | Speech Milestone Q11: Age combined words: Max. Prob. | Speech Milestone Q11: Age combined words: Hazard ratio | SCQ-Q01: Using phrases: Mean | SCQ-Q01: Using phrases: Odds Ratio | SCQ-Q02: Having conversation: Mean | SCQ-Q02: Having conversation: Odds Ratio | SCQ-Q23: Using gestures: Mean | SCQ-Q23: Using gestures: Odds Ratio | SCQ-Q24: Nodding head: Mean | SCQ-Q24: Nodding head: Odds Ratio | SCQ-Q25: Shaking head: Mean | SCQ-Q25: Shaking head: Odds Ratio | SRS-2-Q12: Expressing feelings: Mean | SRS-2-Q12: Expressing feelings: Odds Ratio |
|---|---|---|---|---|---|---|---|---|---|---|---|---|---|---|---|---|---|---|---|---|
| Unaffected siblings: Preschool | | | | | | | | | 1.00 | | 1.00 | | 0.89 | | 1.00 | | 1.00 | | 3.55 | |
| Unaffected siblings: Middle Childhood | | | | | | | | | 1.00 | 1.00 | 0.94 | 0.00 | 0.76 | 0.29 | 0.88 | 0.00 | 1.00 | 1.00 | 3.39 | 0.70 |
| Unaffected siblings: Adolescence | | | | | | | | | 1.00 | 1.00 | 1.00 | 1.00 | 0.50 | 0.15 | 1.00 | 1.00 | 1.00 | 1.00 | 3.09 | **0.35** |
| Controls: Idiopathic ASD: Preschool | 8.15 | | 7.76 | | 0.95 | | 0.86 | | 0.55 | | 0.26 | | 0.49 | | 0.38 | | 0.52 | | 1.96 | |
| Controls: Idiopathic ASD: Middle Childhood | 10.3 | **1.27** | 9.67 | **1.25** | 0.97 | **1.19** | 0.93 | **1.20** | 0.89 | **6.28** | 0.67 | **5.65** | 0.53 | **1.16** | 0.63 | **2.84** | 0.67 | **1.86** | 2.14 | **1.60** |
| Controls: Idiopathic ASD: Adolescence | 11.3 | **1.39** | 10.1 | **1.30** | 0.98 | **1.33** | 0.95 | **1.34** | 0.93 | **10.21** | 0.77 | **9.54** | 0.48 | 0.97 | 0.67 | **3.34** | 0.70 | **2.12** | 2.16 | **1.69** |
| **Discovery: Genetic NDDs (prevalent forms): Preschool** | 6.73 | | 7.03 | | 0.81 | | 0.79 | | 0.45 | | 0.30 | | 0.58 | | 0.52 | | 0.62 | | 2.24 | |
| **Discovery: Genetic NDDs (prevalent forms): Middle Childhood** | 7.87 | **1.17** | 7.82 | 1.11 | 0.90 | 1.17 | 0.84 | 1.05 | 0.72 | **3.13** | 0.59 | **3.34** | 0.59 | 1.03 | 0.66 | **1.77** | 0.68 | 1.30 | 2.24 | 1.00 |
| **Discovery: Genetic NDDs (prevalent forms): Adolescence** | 9.23 | **1.38** | 8.29 | **1.18** | 0.85 | 1.05 | 0.80 | 1.02 | 0.75 | **3.63** | 0.67 | **4.62** | 0.47 | 0.65 | 0.59 | 1.33 | 0.64 | 1.07 | 2.23 | 0.99 |
| Confirmation-1: Genetic NDDs (rare forms): Preschool | 7.49 | | 7.69 | | 0.74 | | 0.76 | | 0.52 | | 0.33 | | 0.58 | | 0.55 | | 0.66 | | 2.21 | |
| Confirmation-1: Genetic NDDs (rare forms): Middle Childhood | 8.08 | 1.08 | 7.54 | 0.98 | 0.85 | 1.14 | 0.79 | 1.04 | 0.70 | **2.13** | 0.56 | **2.54** | 0.59 | 1.02 | 0.58 | 1.14 | 0.64 | 0.93 | 2.38 | 1.45 |
| Confirmation-1: Genetic NDDs (rare forms): Adolescence | 9.68 | **1.30** | 8.42 | 1.11 | 0.89 | 1.33 | 0.82 | 1.12 | 0.82 | **4.08** | 0.66 | **4.12** | 0.52 | 0.79 | 0.59 | 1.22 | 0.63 | 0.88 | 2.29 | 1.22 |
| Confirmation-2: Genetic ASD forms: Preschool | 6.64 | | 6.29 | | 0.85 | | 0.73 | | 0.44 | | 0.20 | | 0.47 | | 0.32 | | 0.48 | | 1.79 | |
| Confirmation-2: Genetic ASD forms: Middle Childhood | 8.88 | **1.34** | 8.41 | **1.34** | 0.90 | 1.26 | 0.81 | 1.20 | 0.78 | **4.41** | 0.58 | **5.42** | 0.54 | 1.30 | 0.54 | **2.47** | 0.59 | **1.57** | 1.98 | 1.58 |
| Confirmation-2: Genetic ASD forms: Adolescence | 9.46 | **1.43** | 8.35 | **1.33** | 0.95 | 1.31 | 0.88 | 1.36 | 0.86 | **7.66** | 0.67 | **7.89** | 0.47 | 0.99 | 0.56 | **2.63** | 0.60 | 1.58 | 2.18 | 2.29 |
| Monogenic ASD forms (Confirmation-2 subset): Preschool | 6.16 | | 5.96 | | 0.81 | | 0.65 | | 0.40 | | 0.19 | | 0.45 | | 0.33 | | 0.48 | | 1.68 | |
| Monogenic ASD forms (Confirmation-2 subset): Middle Childhood | 8.44 | **1.37** | 8.10 | **1.36** | 0.88 | 1.25 | 0.78 | 1.16 | 0.72 | **3.92** | 0.53 | **4.78** | 0.51 | 1.28 | 0.50 | **2.02** | 0.56 | 1.39 | 1.97 | 2.10 |
| Monogenic ASD forms (Confirmation-2 subset): Adolescence | 8.90 | **1.45** | 7.81 | **1.31** | 0.96 | 1.38 | 0.87 | 1.35 | 0.83 | **7.43** | 0.65 | **7.76** | 0.46 | 1.06 | 0.53 | **2.34** | 0.56 | 1.36 | 2.18 | 3.12 |
| CNV-based ASD forms (Confirmation-2 subset): Preschool | 7.85 | | 7.13 | | 0.92 | | 0.85 | | 0.53 | | 0.22 | | 0.52 | | 0.32 | | 0.49 | | 1.95 | |
| CNV-based ASD forms (Confirmation-2 subset): Middle Childhood | 10.1 | **1.29** | 9.27 | 1.30 | 0.96 | 1.43 | 0.89 | 1.42 | 0.88 | **7.20** | 0.67 | **7.42** | 0.59 | 1.37 | 0.63 | **3.92** | 0.66 | **2.07** | 2.00 | 1.08 |
| CNV-based ASD forms (Confirmation-2 subset): Adolescence | 10.9 | **1.38** | 9.72 | **1.36** | 0.94 | 1.20 | 0.92 | 1.49 | 0.91 | **8.83** | 0.70 | **8.16** | 0.49 | 0.87 | 0.61 | **3.32** | 0.67 | 2.13 | 2.20 | 1.44 |
| 1q21.1 deletion: Preschool | 12.7 | | 12.1 | | 0.96 | | 0.94 | | 0.89 | | 0.64 | | 0.57 | | 0.86 | | 0.86 | | 2.52 | |
| 1q21.1 deletion: Middle Childhood | 12.2 | 0.96 | 11.9 | 0.98 | 0.92 | 0.94 | 1.00 | 0.72 | 0.93 | 1.70 | 0.86 | 3.73 | 0.86 | 4.48 | 0.86 | 1.05 | 0.86 | 1.05 | 2.56 | 1.09 |
| 1q21.1 deletion: Adolescence | 13.2 | 1.06 | 11.5 | 0.97 | 1.00 | | 1.00 | | 1.00 | | 0.75 | | 0.75 | | 0.75 | | 1.00 | | 2.29 | 0.59 |
| 16p11.2 duplication: Preschool | 10.8 | | 10.2 | | 0.89 | | 0.91 | | 0.83 | | 0.58 | | 0.42 | | 0.62 | | 0.79 | | 2.39 | |
| 16p11.2 duplication: Middle Childhood | 11.1 | 1.03 | 9.79 | 0.96 | 1.00 | 0.97 | 1.00 | 0.71 | 0.95 | 3.56 | 0.80 | 2.89 | 0.64 | 3.01 | 0.89 | 4.57 | 0.89 | 2.11 | 2.51 | 1.50 |
| 16p11.2 duplication: Adolescence | 10.8 | 0.97 | 8.43 | 0.82 | 1.00 | 1.16 | 1.00 | 0.91 | 1.00 | 1.00 | 0.94 | 10.54 | 0.65 | 2.85 | 0.71 | 1.57 | 0.74 | 0.79 | 2.34 | 1.14 |
| 16p11.2 deletion: Preschool | 9.64 | | 10.5 | | 1.00 | | 1.00 | | 0.73 | | 0.59 | | 0.72 | | 0.81 | | 0.86 | | 2.36 | |
| 16p11.2 deletion: Middle Childhood | 11.0 | 1.15 | 11.3 | 1.08 | 1.00 | 0.81 | 0.97 | 0.71 | 0.94 | 5.41 | 0.86 | **4.25** | 0.63 | 0.67 | 0.88 | 1.76 | 0.89 | 1.36 | 2.24 | 0.70 |
| 16p11.2 deletion: Adolescence | 11.9 | **1.24** | 11.6 | 1.11 | 1.00 | 0.84 | 1.00 | 0.80 | 0.96 | 6.85 | 0.87 | 4.57 | 0.52 | 0.40 | 0.76 | 0.75 | 0.78 | 0.58 | 2.18 | 0.65 |
| 1q21.1 duplication: Preschool | 12.2 | | 12.5 | | 1.00 | | 1.00 | | 0.76 | | 0.35 | | 0.59 | | 0.59 | | 0.71 | | 1.78 | |
| 1q21.1 duplication: Middle Childhood | 7.39 | 0.65 | 6.10 | 0.53 | 0.95 | 0.83 | 0.92 | 1.04 | 0.83 | 1.52 | 0.71 | 4.54 | 0.67 | 1.41 | 0.67 | 1.37 | 0.62 | 0.67 | 2.16 | 2.02 |
| 1q21.1 duplication: Adolescence | 11.5 | 0.94 | 10.1 | 0.81 | 1.00 | 0.75 | 1.00 | 1.55 | 1.00 | 1.00 | 1.00 | 1.00 | 0.80 | 2.76 | 0.80 | 1.72 | 0.93 | 2.86 | 2.29 | 2.45 |
| CSNK2A1: Preschool | 8.44 | | 8.89 | | 0.83 | | 0.82 | | 0.68 | | 0.44 | | 0.64 | | 0.72 | | 0.84 | | 2.40 | |
| CSNK2A1: Middle Childhood | 8.96 | 1.25 | 7.86 | 0.99 | 0.94 | 0.70 | 0.89 | 0.59 | 0.80 | 1.63 | 0.75 | 3.77 | 0.65 | 0.62 | 0.70 | 0.55 | 0.65 | 0.26 | 2.12 | 0.41 |
| CSNK2A1: Adolescence | 10.0 | 1.23 | 8.27 | 0.96 | 1.00 | 1.29 | 1.00 | 1.30 | 1.00 | 1.00 | 0.94 | 18.82 | 0.56 | 0.69 | 0.88 | 2.62 | 0.81 | 0.61 | 2.57 | 1.58 |
| SETBP1: Preschool | 5.33 | | 8.22 | | 0.87 | | 0.73 | | 0.45 | | 0.27 | | 0.82 | | 0.64 | | 0.73 | | 2.60 | |
| SETBP1: Middle Childhood | 6.89 | 1.24 | 7.04 | 0.84 | 0.93 | 1.09 | 0.85 | 0.82 | 0.71 | 2.97 | 0.59 | 3.63 | 0.76 | 0.78 | 0.82 | 2.41 | 0.94 | 6.05 | 2.09 | 0.09 |
| SETBP1: Adolescence | 9.64 | 1.81 | 10.0 | 1.22 | 1.00 | 1.80 | 0.80 | 1.08 | 0.33 | | 0.33 | | 0.67 | | 0.67 | | 0.67 | | 2.00 | 0.25 |
| CTNNB1: Preschool | 7.30 | | 8.79 | | 0.81 | | 0.69 | | 0.51 | | 0.29 | | 0.76 | | 0.61 | | 0.68 | | 2.50 | |
| CTNNB1: Middle Childhood | 9.12 | 1.22 | 8.74 | 0.99 | 0.88 | 0.98 | 0.89 | 0.99 | 0.86 | 6.21 | 0.68 | 5.19 | 0.64 | 0.57 | 0.68 | 1.37 | 0.64 | 0.81 | 2.31 | 1.09 |
| CTNNB1: Adolescence | 8.59 | 1.16 | 8.24 | 0.94 | 0.83 | 0.64 | 0.64 | 0.56 | 0.69 | 2.16 | 0.62 | 3.85 | 0.54 | 0.38 | 0.69 | 1.59 | 0.69 | 1.09 | 2.45 | 1.12 |
| SLC6A1: Preschool | 6.22 | | 5.22 | | 1.00 | | 0.86 | | 0.58 | | 0.42 | | 0.65 | | 0.54 | | 0.54 | | 2.30 | |
| SLC6A1: Middle Childhood | 9.12 | 1.46 | 8.14 | 1.56 | 0.91 | 1.65 | 0.80 | 0.81 | 0.68 | 1.38 | 0.55 | 1.51 | 0.59 | 0.83 | 0.64 | 1.37 | 0.64 | 1.49 | 2.29 | 1.01 |
| SLC6A1: Adolescence | 8.46 | 1.37 | 6.86 | 1.23 | 0.93 | 0.70 | 0.79 | 0.67 | 0.80 | 3.05 | 0.60 | 2.04 | 0.47 | 0.40 | 0.67 | 1.80 | 0.67 | 1.65 | 2.22 | 0.66 |
| MED13L: Preschool | 6.59 | | 7.28 | | 0.78 | | 0.51 | | 0.24 | | 0.17 | | 0.83 | | 0.55 | | 0.59 | | 2.00 | |
| MED13L: Middle Childhood | 5.92 | 0.90 | 7.97 | 1.10 | 0.87 | 0.87 | 0.80 | 0.93 | 0.53 | 5.35 | 0.33 | 3.62 | 0.73 | 0.63 | 0.67 | 2.38 | 0.73 | 2.58 | 2.12 | 1.49 |
| MED13L: Adolescence | 10.1 | 1.54 | 8.57 | 1.17 | 0.90 | 0.77 | 0.75 | 0.65 | 0.67 | 6.00 | 0.67 | 9.16 | 0.50 | 0.19 | 0.67 | 1.43 | 0.83 | 3.34 | 1.43 | 0.14 |
| PPP2R5D: Preschool | 5.45 | | 6.01 | | 0.72 | | 0.80 | | 0.35 | | 0.24 | | 0.62 | | 0.54 | | 0.59 | | 2.57 | |
| PPP2R5D: Middle Childhood | 7.15 | 1.23 | 8.16 | 1.32 | 0.93 | 1.56 | 0.87 | 0.78 | 0.72 | 4.89 | 0.55 | 3.90 | 0.45 | 0.50 | 0.66 | 1.59 | 0.62 | 1.11 | 2.37 | 0.64 |
| PPP2R5D: Adolescence | 6.45 | 1.24 | 6.68 | 1.14 | 0.58 | 0.62 | 0.50 | 0.31 | 0.42 | 1.43 | 0.33 | 1.72 | 0.25 | 0.21 | 0.58 | 1.27 | 0.42 | 0.54 | 2.12 | 0.29 |
| GRIN2B: Preschool | 6.63 | | 5.83 | | 1.00 | | 0.76 | | 0.42 | | 0.23 | | 0.39 | | 0.42 | | 0.48 | | 1.85 | |
| GRIN2B: Middle Childhood | 5.37 | 0.85 | 4.79 | 0.85 | 0.78 | 0.92 | 0.76 | 0.67 | 0.50 | 1.40 | 0.25 | 1.14 | 0.42 | 1.15 | 0.25 | 0.46 | 0.42 | 0.74 | 2.02 | 1.36 |
| GRIN2B: Adolescence | 6.86 | 1.06 | 6.17 | 1.09 | 0.83 | 0.94 | 0.78 | 0.67 | 0.71 | 3.64 | 0.57 | 5.39 | 0.29 | 0.69 | 0.50 | 1.37 | 0.50 | 0.99 | 2.24 | 2.37 |
| SYNGAP1: Preschool | 4.00 | | 4.97 | | 0.52 | | 0.10 | | 0.20 | | 0.12 | | 0.56 | | 0.20 | | 0.52 | | 1.83 | |
| SYNGAP1: Middle Childhood | 3.63 | 0.92 | 5.21 | 1.05 | 0.76 | 1.41 | 0.50 | 1.94 | 0.47 | 3.53 | 0.35 | 4.85 | 0.59 | 1.10 | 0.59 | 5.93 | 0.65 | 1.71 | 1.73 | 0.72 |
| SYNGAP1: Adolescence | 9.56 | 2.52 | 8.56 | 1.72 | 0.64 | 1.08 | 0.55 | 1.97 | 0.67 | 8.93 | 0.50 | 11.81 | 0.33 | 0.36 | 0.33 | 2.42 | 0.75 | 3.14 | 1.95 | 1.38 |
| SCN2A: Preschool | 5.00 | | 4.12 | | 0.51 | | 0.38 | | 0.11 | | 0.02 | | 0.31 | | 0.16 | | 0.33 | | 1.78 | |
| SCN2A: Middle Childhood | 3.00 | 0.60 | 2.94 | 0.71 | 0.75 | 1.14 | 0.65 | 1.79 | 0.43 | 6.33 | 0.22 | 11.44 | 0.38 | 1.32 | 0.27 | 1.88 | 0.38 | 1.11 | 1.89 | 1.28 |
| SCN2A: Adolescence | 6.52 | 1.18 | 6.48 | 1.52 | 0.75 | 1.54 | 0.64 | 1.94 | 0.47 | 6.85 | 0.40 | 29.62 | 0.20 | 0.55 | 0.20 | 1.38 | 0.33 | 1.02 | 2.43 | 4.31 |
| ASXL3: Preschool | 4.50 | | 4.57 | | 0.79 | | 0.43 | | 0.12 | | 0.00 | | 0.50 | | 0.44 | | 0.56 | | 1.17 | |
| ASXL3: Middle Childhood | 4.88 | 0.85 | 5.92 | 1.11 | 0.61 | 0.26 | 0.51 | 0.18 | 0.24 | 2.11 | 0.18 | 1.00 | 0.59 | 1.50 | 0.35 | 0.70 | 0.35 | 0.42 | 2.05 | 10.11 |
| ASXL3: Adolescence | 3.88 | 0.93 | 3.50 | 0.83 | 0.75 | | 0.67 | | 0.29 | 2.16 | 0.29 | 1.00 | 0.29 | 0.43 | 0.29 | 0.32 | 0.29 | 0.17 | 1.64 | 10.08 |
| STXBP1: Preschool | 3.64 | | 3.50 | | 0.49 | | 0.27 | | 0.07 | | 0.07 | | 0.38 | | 0.17 | | 0.40 | | 2.25 | |
| STXBP1: Middle Childhood | 2.20 | 0.57 | 1.84 | **0.49** | 0.78 | 2.28 | 0.60 | 2.16 | 0.26 | 4.86 | 0.19 | 3.12 | 0.41 | 1.11 | 0.26 | 1.79 | 0.33 | 0.73 | 1.81 | 0.48 |
| STXBP1: Adolescence | 2.04 | 0.49 | 1.46 | **0.40** | 0.35 | 0.55 | 0.27 | 0.54 | 0.16 | 2.22 | 0.11 | 1.33 | 0.26 | 0.52 | 0.05 | 0.25 | 0.11 | 0.17 | 1.78 | 0.45 |

log10 (effect size): 1.0, 0.5, 0.0, -0.5, -1.0

**Supplementary Figure 2: Age-stratified developmental trajectories across all testing groups**

Shown are the cross-sectional age-stratified analyses of age at evaluation groups: "Middle Childhood" (6 to 11 years, 11 months) and "Adolescence" (12 to 17 years, 11 months), each tested against "Preschool" (0 to 5 years, 11 months). Age-group-stratified statistical tests were not performed for any testing group with fewer than five measurements. The effect sizes for significant and suggestive association signals are highlighted in red (indicating lower communication outcomes than the "Preschool" group) and blue (indicating higher communication outcomes). Bolded values indicate statistically significant differences between communication outcomes after multiple testing correction; non-bold colored values reflect suggestive differences. Legend: Max Prob.: maximum cumulative probability of milestone attainment at the latest observed follow-up; SCQ: Social Communication Questionnaire; SRS-2: Social Responsiveness Scale, Second Edition.

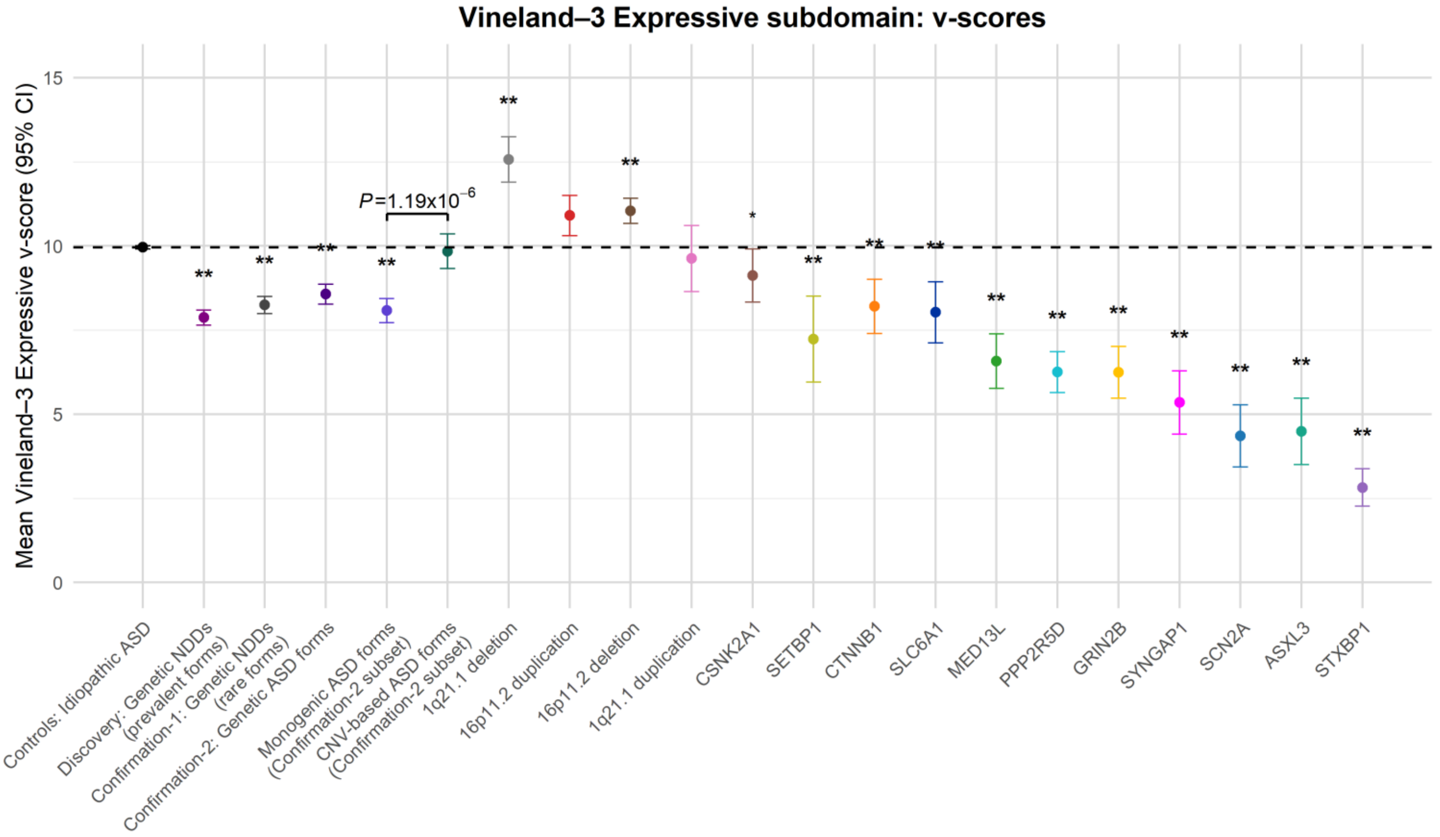


**Supplementary Figure 3: Association analyses for Vineland-3 Expressive subdomain**

Shown are the group-level means and t-based 95% confidence intervals (CIs) of the Vineland-3 Expressive subdomain (v-scores) for individuals in the 1) idiopathic ASD control, 2) *Discovery*, 3) *Confirmation-1*, and 4) *Confirmation-2* cohorts, followed by two subsets of the *Confirmation-2* cohort: 5) Monogenic ASD forms and 6) CNV-based ASD forms, and 7-21) 15 genetic conditions represented in the *Discovery* cohort. The black dotted horizontal line indicates the mean Expressive v-score of the idiopathic ASD control group, which served as the reference for all case-control comparisons. Statistical comparisons between each case group and the idiopathic ASD controls were conducted using Gamma generalised linear models (Gamma-GLM) with a log link function, adjusting for sex. Asterisks denote statistically significant group differences after Bonferroni multiple testing correction: * $P < 0.016$ (suggestive), ** $P < 7.94\times10^{-4}$ (significant). The single displayed *P*-value reflects the comparison between monogenic and CNV-based ASD forms within the Confirmation-2 cohort.

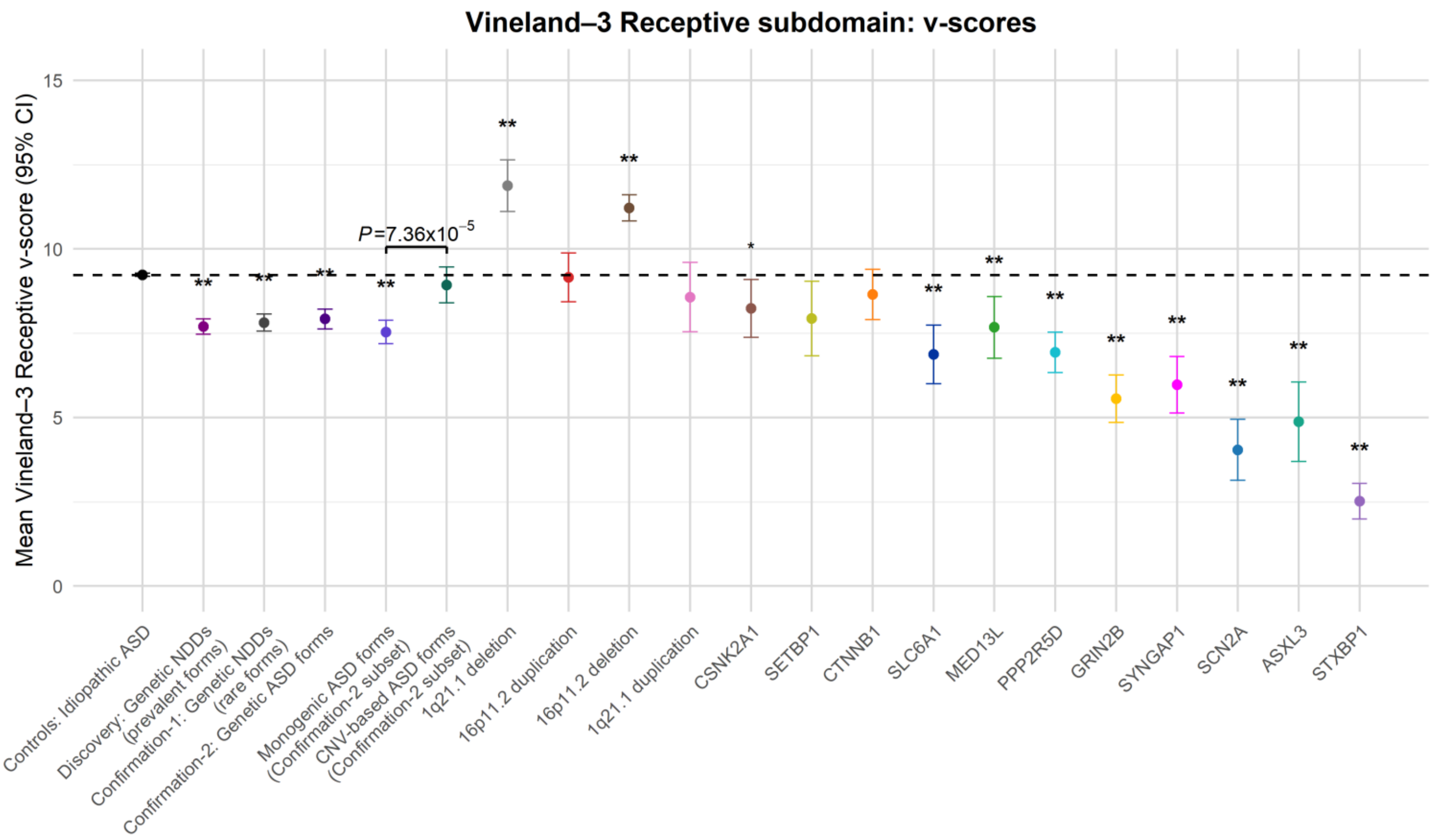


**Supplementary Figure 4: Association analyses for Vineland-3 Receptive subdomain**

Shown are the group-level means and t-based 95% confidence intervals (CIs) of the Vineland-3 Receptive subdomain (v-scores) for individuals in the 1) idiopathic ASD control, 2) *Discovery*, 3) *Confirmation-1*, and 4) *Confirmation-2* cohorts, followed by two subsets of the *Confirmation-2* cohort: 5) Monogenic ASD forms and 6) CNV-based ASD forms, and 7-21) 15 genetic conditions represented in the *Discovery* cohort. The black dotted horizontal line indicates the mean Receptive v-score of the idiopathic ASD control group, which served as the reference for all case-control comparisons. Statistical comparisons between each case group and the idiopathic ASD controls were conducted using Gamma generalised linear models (Gamma-GLM) with a log link function, adjusting for sex. Asterisks denote statistically significant group differences after Bonferroni multiple testing correction: * $P < 0.016$ (suggestive), ** $P < 7.94\times10^{-4}$ (significant). The single displayed *P*-value reflects the comparison between monogenic and CNV-based ASD forms within the Confirmation-2 cohort.

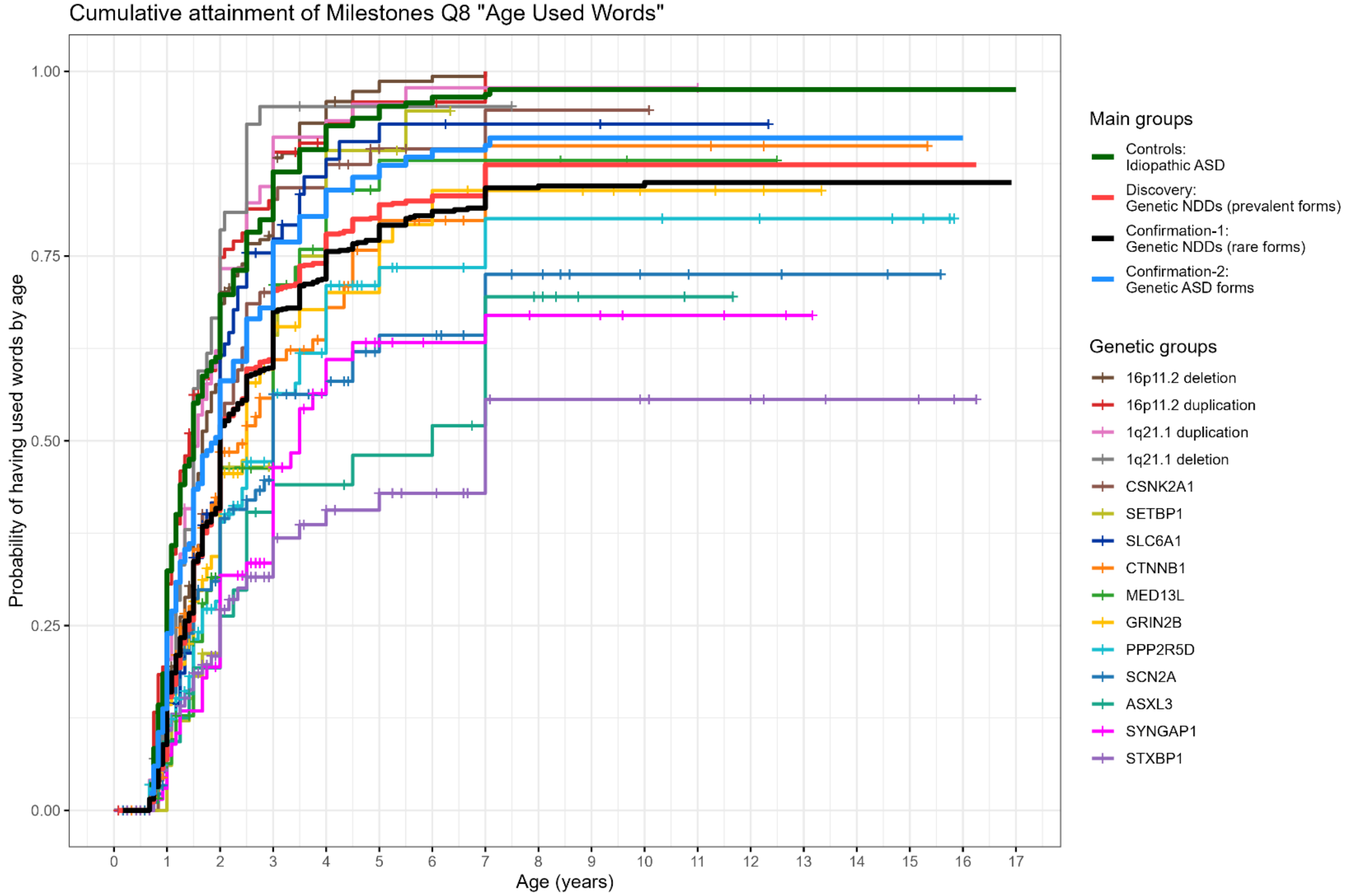


**Supplementary Figure 5: Cumulative attainment of speech milestone Q8: "Age Used Words" across testing groups**

Shown are the Kaplan-Meier cumulative probability curves for the speech milestone "Age Used Words" (Speech milestone Q8), representing the probability of having used single words by a given age across study groups. Plotted are the idiopathic ASD, *Discovery*, *Confirmation-1*, and *Confirmation-2* cohorts, and the 15 genetic NDD conditions present in the Discovery cohort. Censoring marks (+) indicate participants who had not yet attained the milestone by the age of evaluation. Statistical differences between curves were evaluated using Cox proportional hazards regression models adjusted for sex and are detailed in Supplementary Table 2.

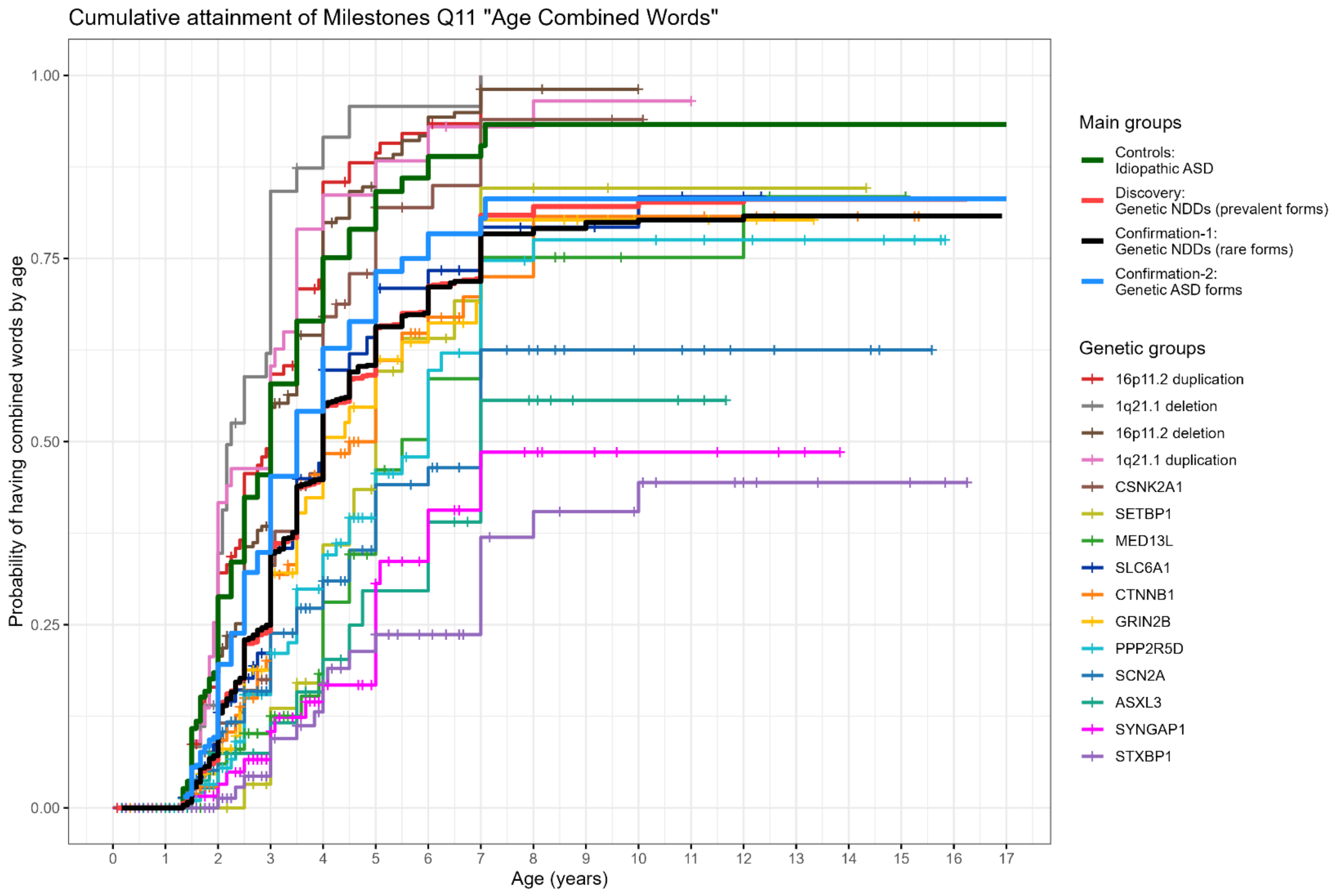


**Supplementary Figure 6: Cumulative attainment of speech milestone Q11: "Age Combined Words" across testing groups**

Shown are the Kaplan-Meier cumulative probability curves for the speech milestone "Age Combined Words" (Speech milestone Q11), representing the probability of having combined single words by a given age across study groups. Plotted are the idiopathic ASD, *Discovery*, *Confirmation-1*, and *Confirmation-2* cohorts, and the 15 genetic NDD conditions present in the Discovery cohort. Censoring marks (+) indicate participants who had not yet attained the milestone by the age of evaluation. Statistical differences between curves were evaluated using Cox proportional hazards regression models adjusted for sex and are detailed in Supplementary Table 2.

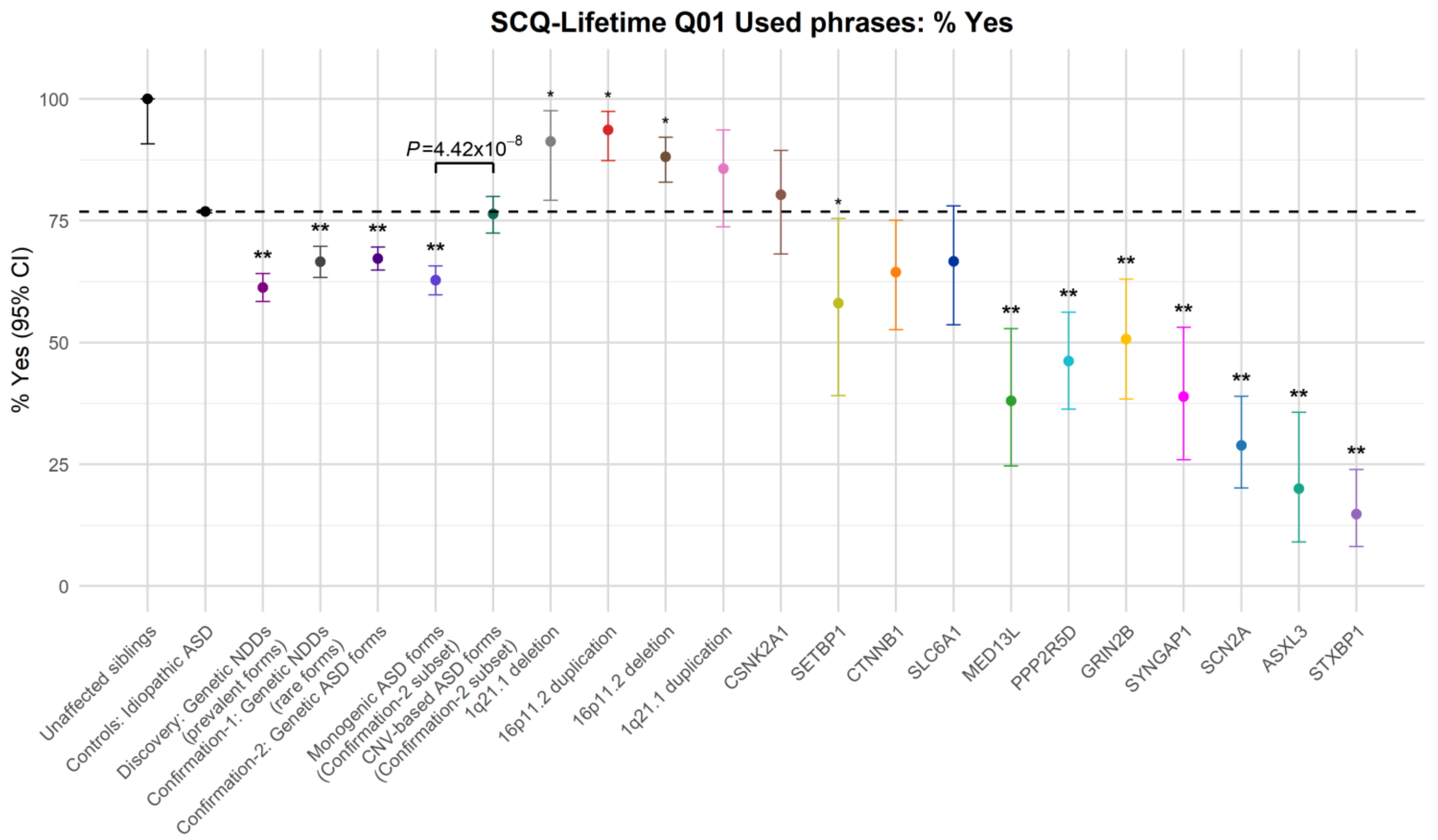


**Supplementary Figure 7: Association analyses for SCQ Q01 "Using phrases"**

Shown are the group-level means and Clopper-Pearson 95% confidence intervals (CIs) of the SCQ Q01 "Using phrases" (percent "Yes") outcome for individuals in the 1) unaffected siblings, 2) idiopathic ASD control, 3) *Discovery*, 4) *Confirmation-1*, and 5) *Confirmation-2* cohorts, followed by two subsets of the *Confirmation-2* cohort: 6) Monogenic ASD forms and 7) CNV-based ASD forms, and 8-22) 15 genetic conditions represented in the *Discovery* cohort. The "unaffected siblings" group is plotted only for reference and was not included in statistical testing. The black dotted horizontal line indicates the mean SCQ Q01 outcome of the idiopathic ASD control group, which served as the reference for all case-control comparisons. Statistical comparisons between each case group and the idiopathic ASD controls were conducted using binomial logistic regression with a logit link adjusted for sex and age at evaluation. Asterisks denote statistically significant group differences after Bonferroni multiple testing correction: * $P < 0.016$ (suggestive), ** $P < 7.94\times10^{-4}$ (significant). The single displayed $P$-value reflects the comparison between monogenic and CNV-based ASD forms within the Confirmation-2 cohort.

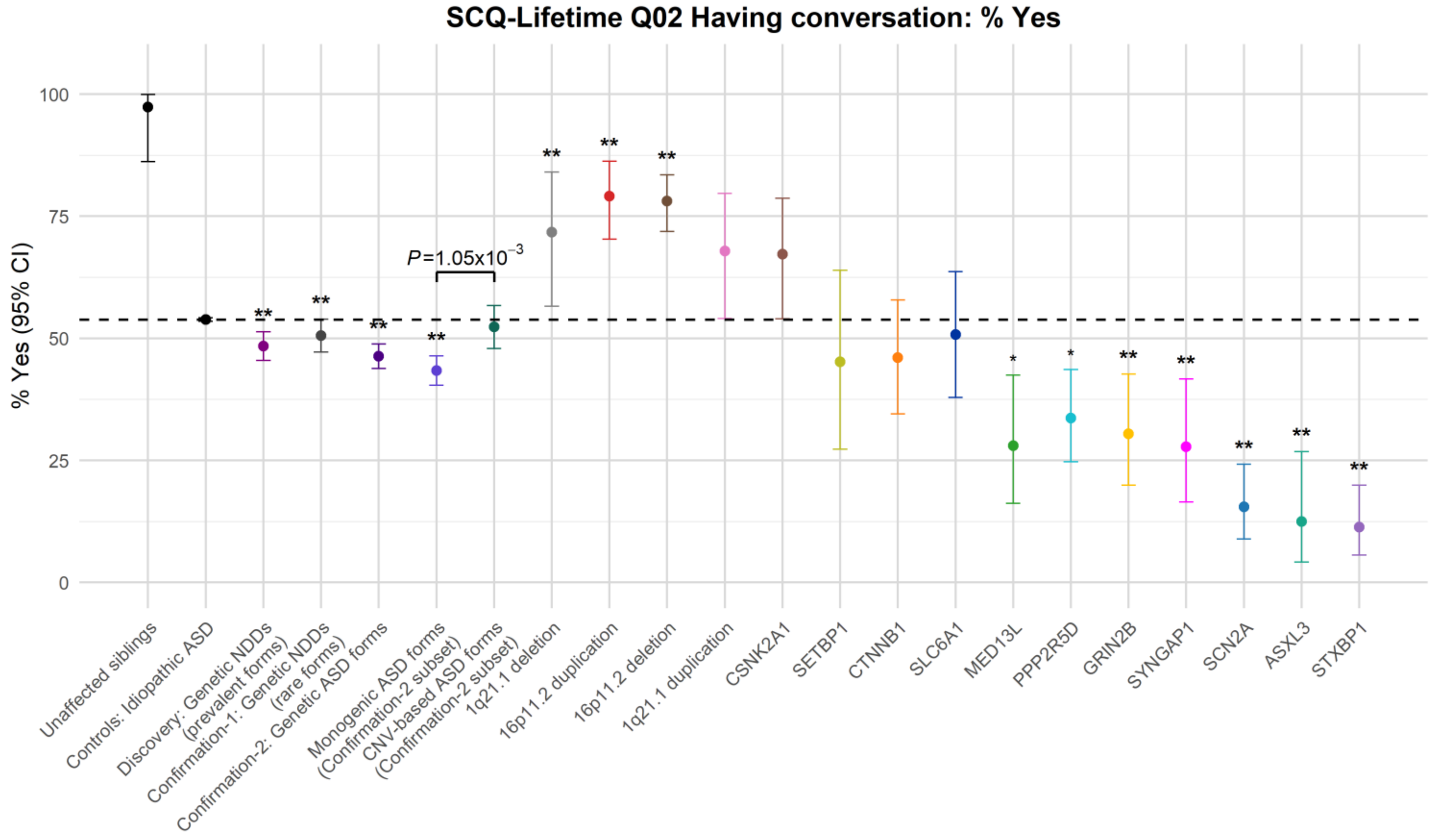


**Supplementary Figure 8: Association analyses for SCQ Q02 "Having conversation"**

Shown are the group-level means and Clopper-Pearson 95% confidence intervals (CIs) of the SCQ Q02 "Having conversation" (percent "Yes") outcome for individuals in the 1) unaffected siblings, 2) idiopathic ASD control, 3) *Discovery*, 4) *Confirmation-1*, and 5) *Confirmation-2* cohorts, followed by two subsets of the *Confirmation-2* cohort: 6) Monogenic ASD forms and 7) CNV-based ASD forms, and 8-22) 15 genetic conditions represented in the *Discovery* cohort. The "unaffected siblings" group is plotted only for reference and was not included in statistical testing. The black dotted horizontal line indicates the mean SCQ Q02 outcome of the idiopathic ASD control group, which served as the reference for all case-control comparisons. Statistical comparisons between each case group and the idiopathic ASD controls were conducted using binomial logistic regression with a logit link adjusted for sex and age at evaluation. Asterisks denote statistically significant group differences after Bonferroni multiple testing correction: * $P < 0.016$ (suggestive), ** $P < 7.94\times10^{-4}$ (significant). The single displayed *P*-value reflects the comparison between monogenic and CNV-based ASD forms within the Confirmation-2 cohort.

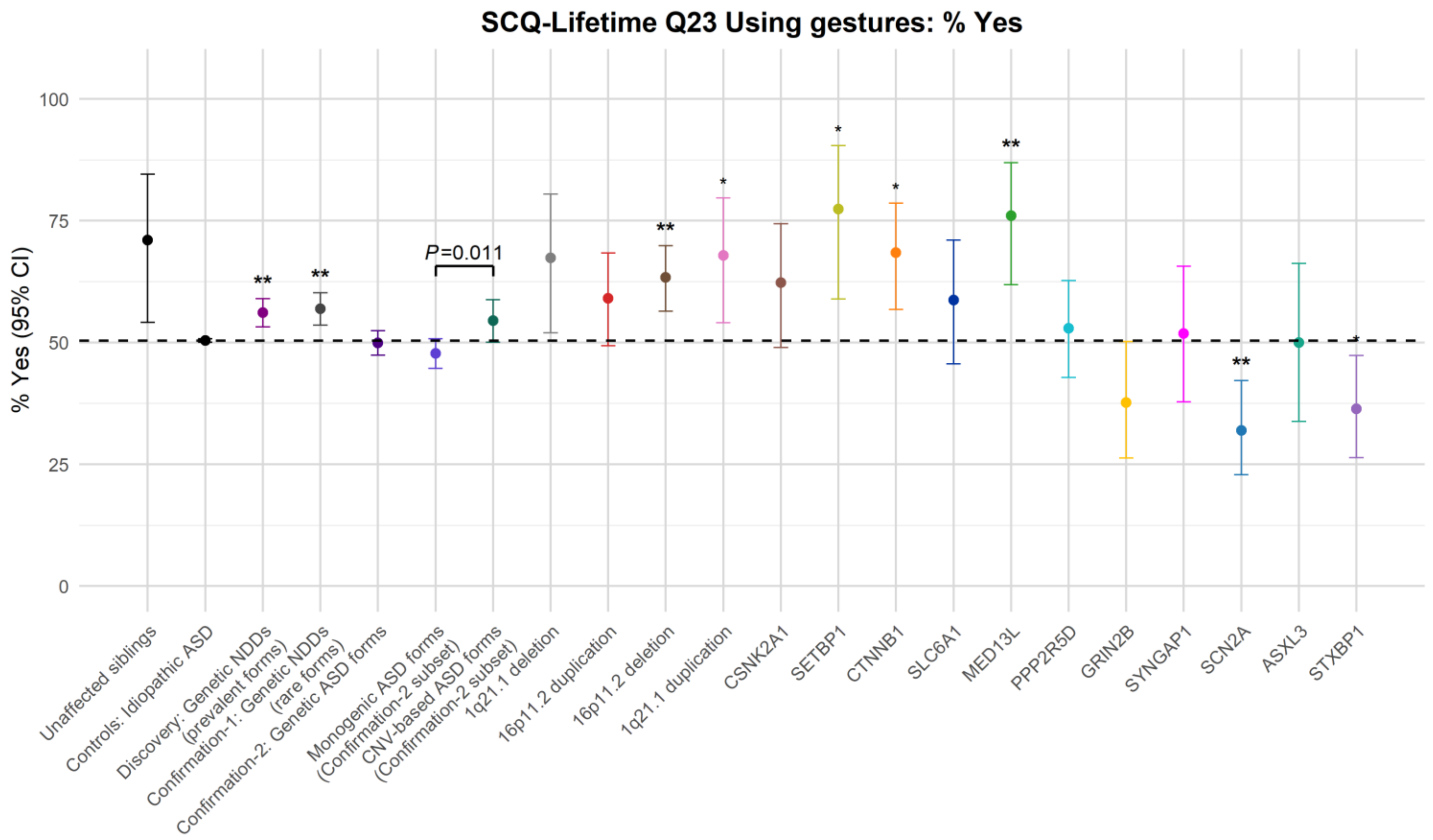


**Supplementary Figure 9: Association analyses for SCQ Q23 "Using gestures"**

Shown are the group-level means and Clopper-Pearson 95% confidence intervals (CIs) of the SCQ Q23 "Using gestures" (percent "Yes") outcome for individuals in the 1) unaffected siblings, 2) idiopathic ASD control, 3) *Discovery*, 4) *Confirmation-1*, and 5) *Confirmation-2* cohorts, followed by two subsets of the *Confirmation-2* cohort: 6) Monogenic ASD forms and 7) CNV-based ASD forms, and 8-22) 15 genetic conditions represented in the *Discovery* cohort. The "unaffected siblings" group is plotted only for reference and was not included in statistical testing. The black dotted horizontal line indicates the mean SCQ Q23 outcome of the idiopathic ASD control group, which served as the reference for all case-control comparisons. Statistical comparisons between each case group and the idiopathic ASD controls were conducted using binomial logistic regression with a logit link adjusted for sex and age at evaluation. Asterisks denote statistically significant group differences after Bonferroni multiple testing correction: * $P < 0.016$ (suggestive), ** $P < 7.94\times10^{-4}$ (significant). The single displayed $P$-value reflects the comparison between monogenic and CNV-based ASD forms within the Confirmation-2 cohort.

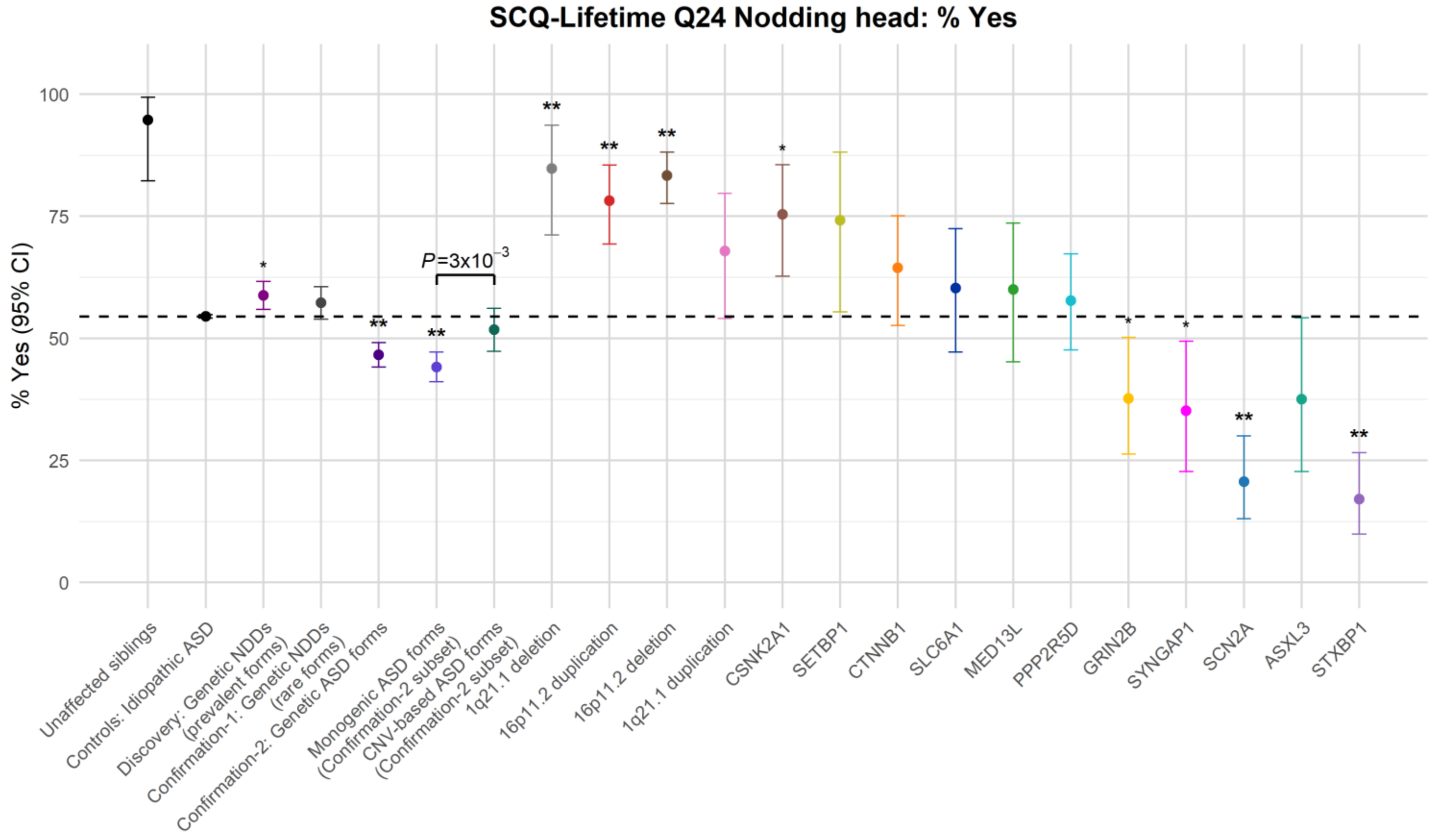


**Supplementary Figure 10: Association analyses for SCQ Q24 "Nodding head"**

Shown are the group-level means and Clopper-Pearson 95% confidence intervals (CIs) of the SCQ Q24 "Nodding head" (percent "Yes") outcome for individuals in the 1) unaffected siblings, 2) idiopathic ASD control, 3) *Discovery*, 4) *Confirmation-1*, and 5) *Confirmation-2* cohorts, followed by two subsets of the *Confirmation-2* cohort: 6) Monogenic ASD forms and 7) CNV-based ASD forms, and 8-22) 15 genetic conditions represented in the *Discovery* cohort. The "unaffected siblings" group is plotted only for reference and was not included in statistical testing. The black dotted horizontal line indicates the mean SCQ Q24 outcome of the idiopathic ASD control group, which served as the reference for all case-control comparisons. Statistical comparisons between each case group and the idiopathic ASD controls were conducted using binomial logistic regression with a logit link adjusted for sex and age at evaluation. Asterisks denote statistically significant group differences after Bonferroni multiple testing correction: * $P < 0.016$ (suggestive), ** $P < 7.94\times10^{-4}$ (significant). The single displayed $P$-value reflects the comparison between monogenic and CNV-based ASD forms within the Confirmation-2 cohort.

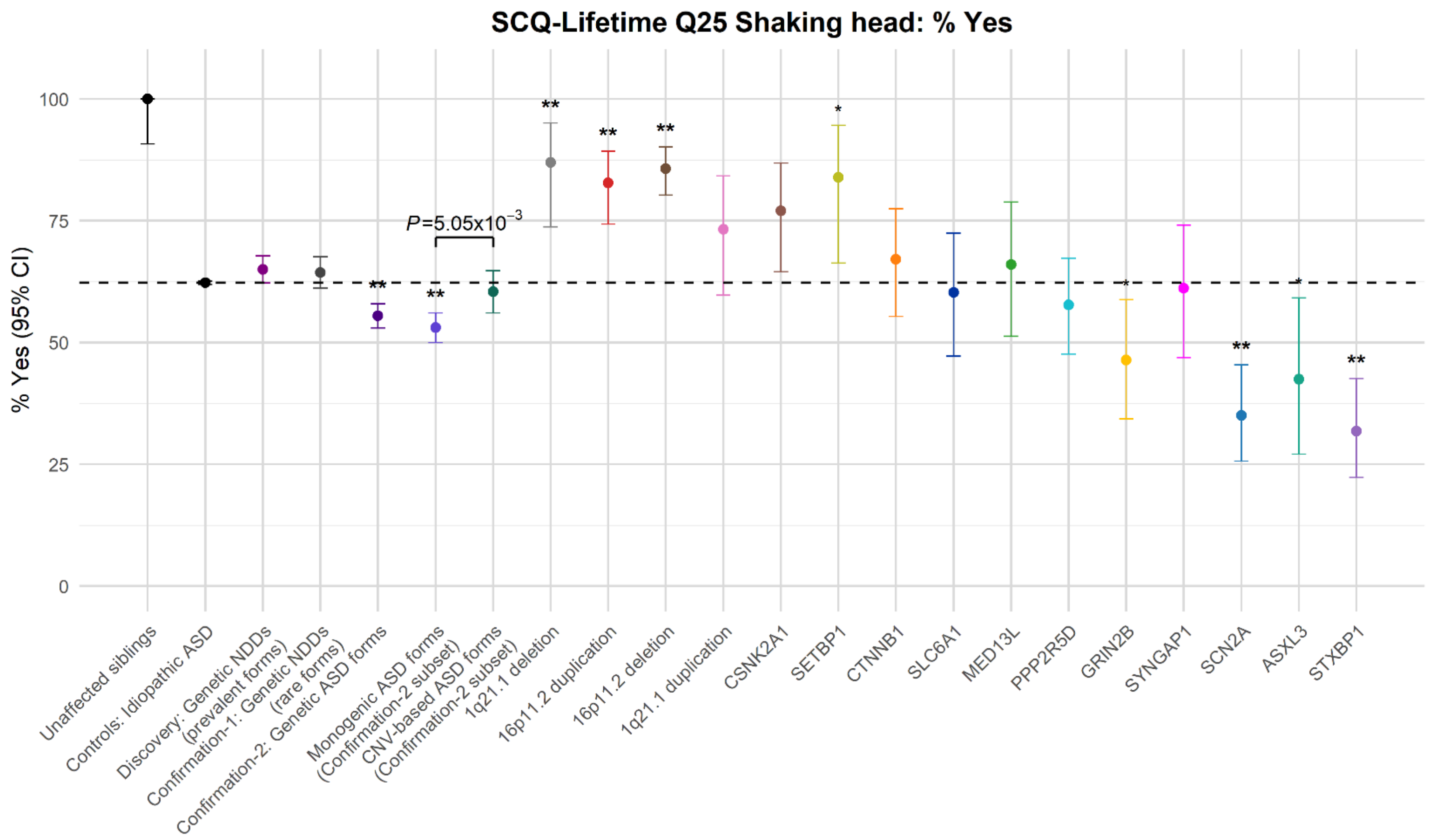


**Supplementary Figure 11: Association analyses for SCQ Q25 "Shaking head"**

Shown are the group-level means and Clopper-Pearson 95% confidence intervals (CIs) of the SCQ Q25 "Shaking head" (percent "Yes") outcome for individuals in the 1) unaffected siblings, 2) idiopathic ASD control, 3) *Discovery*, 4) *Confirmation-1*, and 5) *Confirmation-2* cohorts, followed by two subsets of the *Confirmation-2* cohort: 6) Monogenic ASD forms and 7) CNV-based ASD forms, and 8-22) 15 genetic conditions represented in the *Discovery* cohort. The "unaffected siblings" group is plotted only for reference and was not included in statistical testing. The black dotted horizontal line indicates the mean SCQ Q25 outcome of the idiopathic ASD control group, which served as the reference for all case-control comparisons. Statistical comparisons between each case group and the idiopathic ASD controls were conducted using binomial logistic regression with a logit link adjusted for sex and age at evaluation. Asterisks denote statistically significant group differences after Bonferroni multiple testing correction: * $P < 0.016$ (suggestive), ** $P < 7.94\times10^{-4}$ (significant). The single displayed $P$-value reflects the comparison between monogenic and CNV-based ASD forms within the Confirmation-2 cohort.

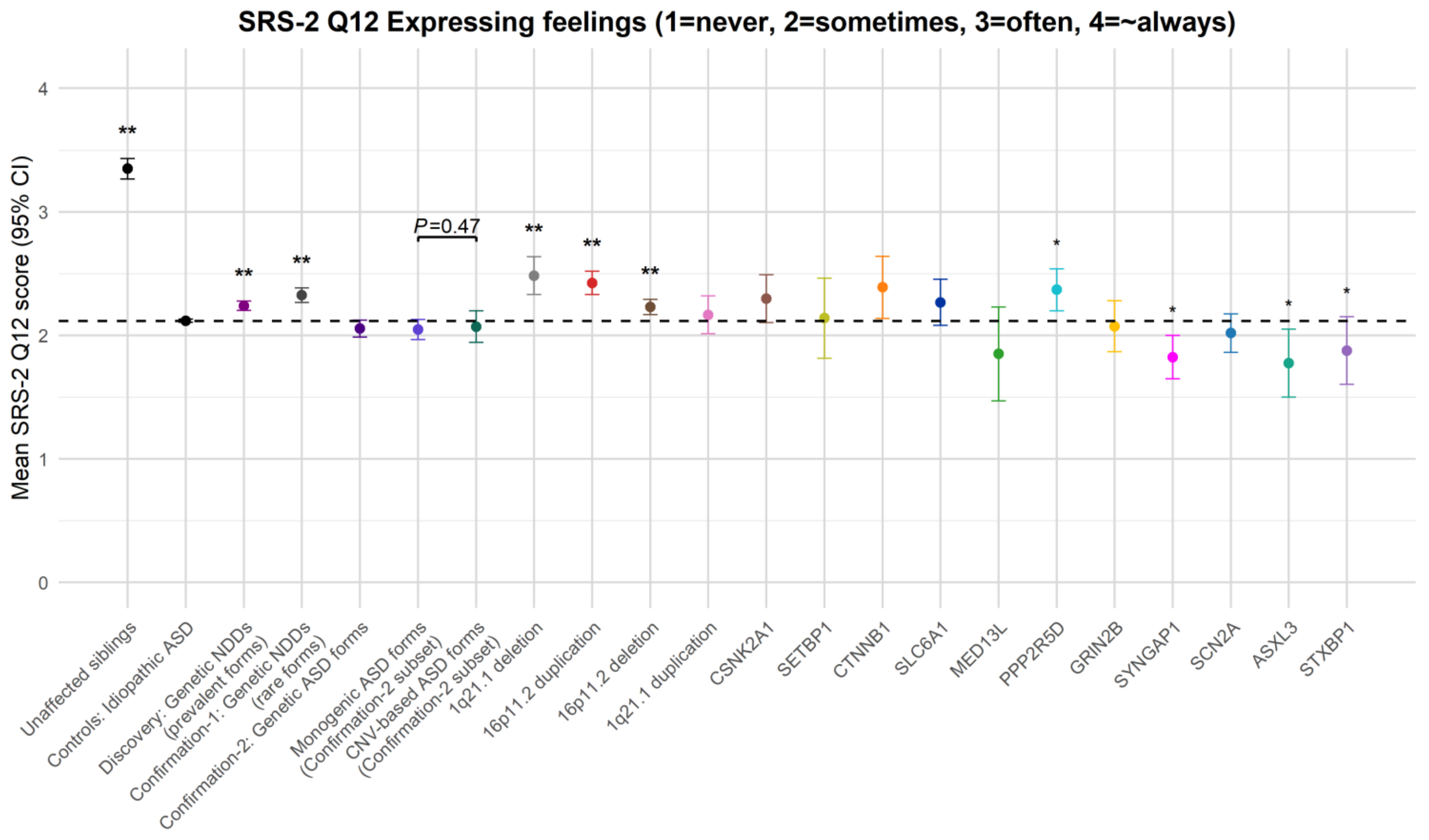


**Supplementary Figure 12: Association analyses for SRS-2 Q12 "Expressing feelings"**

Shown are the group-level means and t-based 95% confidence intervals (CIs) of the SRS-2 Q12 "Expressing feelings" (ordinal: 1 = Not true; 2 = Sometimes true; 3 = Often true; 4 = Almost always true) outcome for individuals in the 1) unaffected siblings, 2) idiopathic ASD control, 3) *Discovery*, 4) *Confirmation-1*, and 5) *Confirmation-2* cohorts, followed by two subsets of the *Confirmation-2* cohort: 6) Monogenic ASD forms and 7) CNV-based ASD forms, and 8-22) 15 genetic conditions represented in the *Discovery* cohort. The black dotted horizontal line indicates the mean SRS-2 Q12 outcome of the idiopathic ASD control group, which served as the reference for all case-control comparisons. Statistical comparisons between each case group and the idiopathic ASD controls were conducted using proportional odds (cumulative logit) regression adjusted for sex and age at evaluation. Asterisks denote statistically significant group differences after Bonferroni multiple testing correction: * $P < 0.015$ (suggestive), ** $P < 7.58\times10^{-4}$ (significant). The single displayed *P*-value reflects the comparison between monogenic and CNV-based ASD forms within the Confirmation-2 cohort.

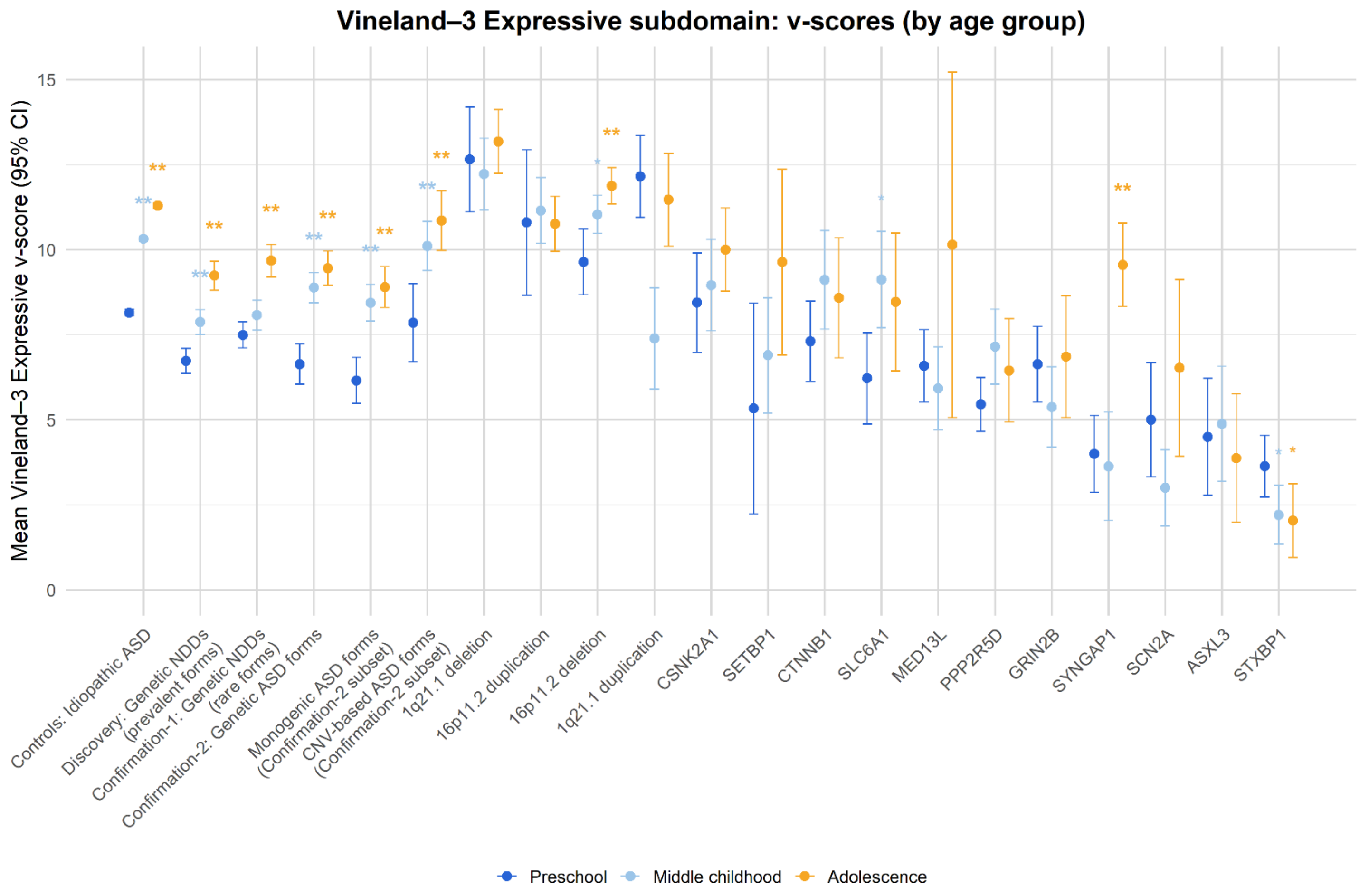


**Supplementary Figure 13: Age-stratified developmental trajectories for Vineland-3 Expressive subdomain** Shown are the group-level means and t-based 95% confidence intervals (CIs) of the Vineland-3 Expressive subdomain (v-scores), stratified by age at evaluation: Preschool (0 to 5 years, 11 months; dark blue), Middle Childhood (6 to 11 years, 11 months; light blue), and Adolescence (12 to 17 years, 11 months; orange). Each age group is plotted separately for the 1) idiopathic ASD control, 2) *Discovery*, 3) *Confirmation-1*, and 4) *Confirmation-2* cohorts, followed by two subsets of the *Confirmation-2* cohort: 5) Monogenic ASD forms and 6) CNV-based ASD forms, and 7-21) 15 genetic conditions represented in the *Discovery* cohort. Within each genetic group, statistical comparisons were performed between Preschool (reference) and each older age group using Gamma generalised linear models (Gamma-GLM) with a log link, adjusting for sex. Asterisks denote statistically significant group differences after Bonferroni multiple testing correction: * $P < 0.016$ (suggestive), ** $P < 7.94\times10^{-4}$ (significant).

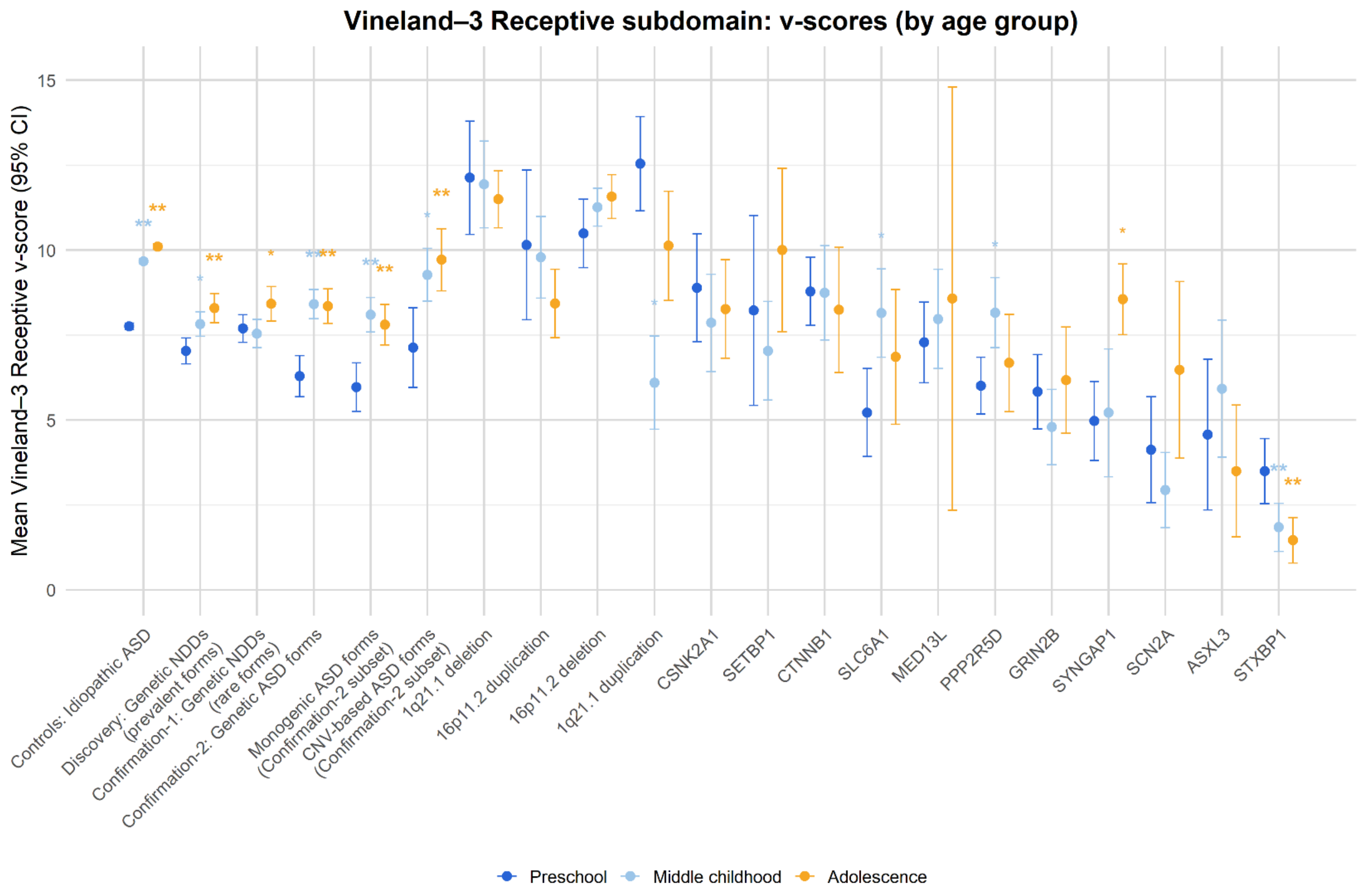


**Supplementary Figure 14: Age-stratified developmental trajectories for Vineland-3 Receptive subdomain** Shown are the group-level means and t-based 95% confidence intervals (CIs) of the Vineland-3 Receptive subdomain (v-scores), stratified by age at evaluation: Preschool (0 to 5 years, 11 months; dark blue), Middle Childhood (6 to 11 years, 11 months; light blue), and Adolescence (12 to 17 years, 11 months; orange). Each age group is plotted separately for the 1) idiopathic ASD control, 2) *Discovery*, 3) *Confirmation-1*, and 4) *Confirmation-2* cohorts, followed by two subsets of the *Confirmation-2* cohort: 5) Monogenic ASD forms and 6) CNV-based ASD forms, and 7-21) 15 genetic conditions represented in the *Discovery* cohort. Within each genetic group, statistical comparisons were performed between Preschool (reference) and each older age group using Gamma generalised linear models (Gamma-GLM) with a log link, adjusting for sex. Asterisks denote statistically significant group differences after Bonferroni multiple testing correction: * $P < 0.016$ (suggestive), ** $P < 7.94\times10^{-4}$ (significant).

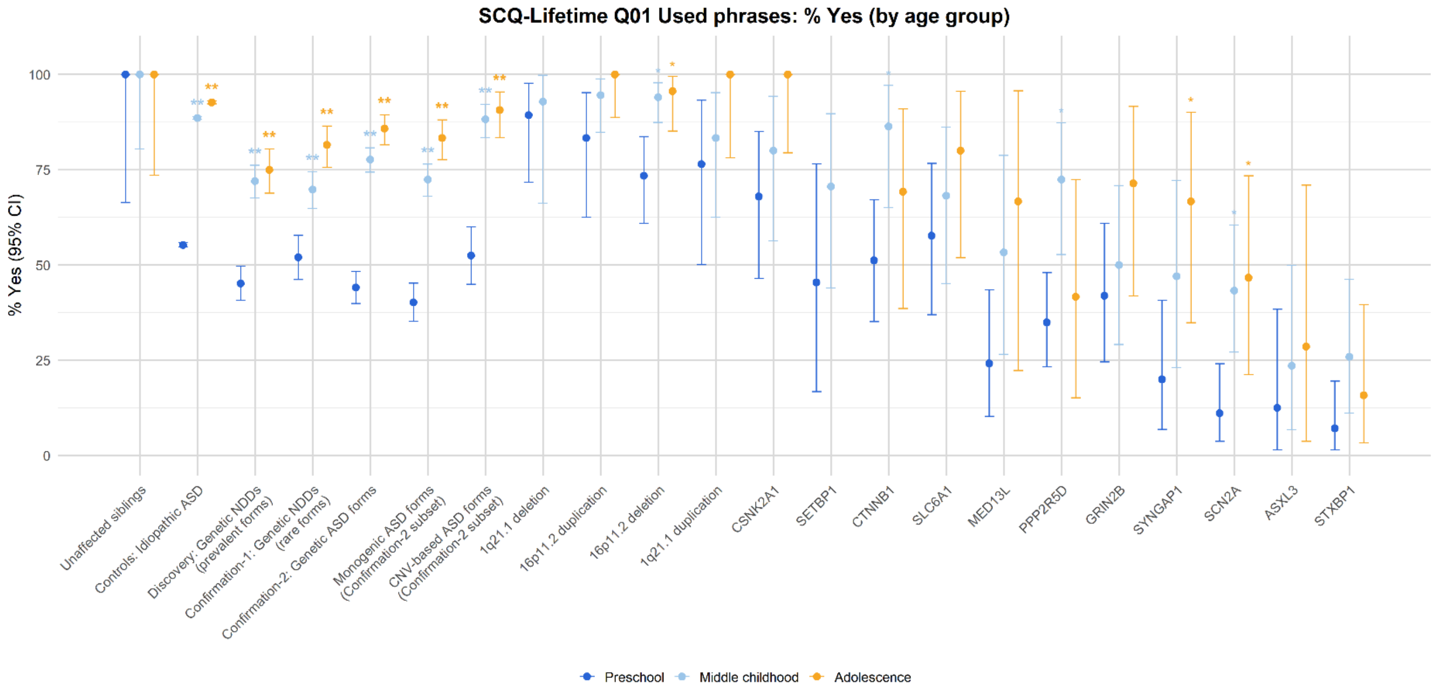


**Supplementary Figure 15: Age-stratified developmental trajectories for SCQ Q01 "Using phrases"**

Shown are the group-level means and Clopper-Pearson 95% confidence intervals (CIs) of the SCQ Q01 "Using phrases" (percent "Yes") outcome, stratified by age at evaluation: Preschool (0 to 5 years, 11 months; dark blue), Middle Childhood (6 to 11 years, 11 months; light blue), and Adolescence (12 to 17 years, 11 months; orange). Each age group is plotted separately for the 1) Unaffected siblings, 2) idiopathic ASD control, 3) *Discovery*, 4) *Confirmation-1*, and 5) *Confirmation-2* cohorts, followed by two subsets of the *Confirmation-2* cohort: 6) Monogenic ASD forms and 7) CNV-based ASD forms, and 8-22) 15 genetic conditions represented in the *Discovery* cohort. Within each genetic group, statistical comparisons were performed between Preschool (reference) and each older age group using binomial logistic regression with a logit link adjusted for sex and age at evaluation. Groups with fewer than five measurements were excluded from statistical testing and are not shown. Asterisks denote statistically significant group differences after Bonferroni multiple testing correction: * $P < 0.016$ (suggestive), ** $P < 7.94\times10^{-4}$ (significant).

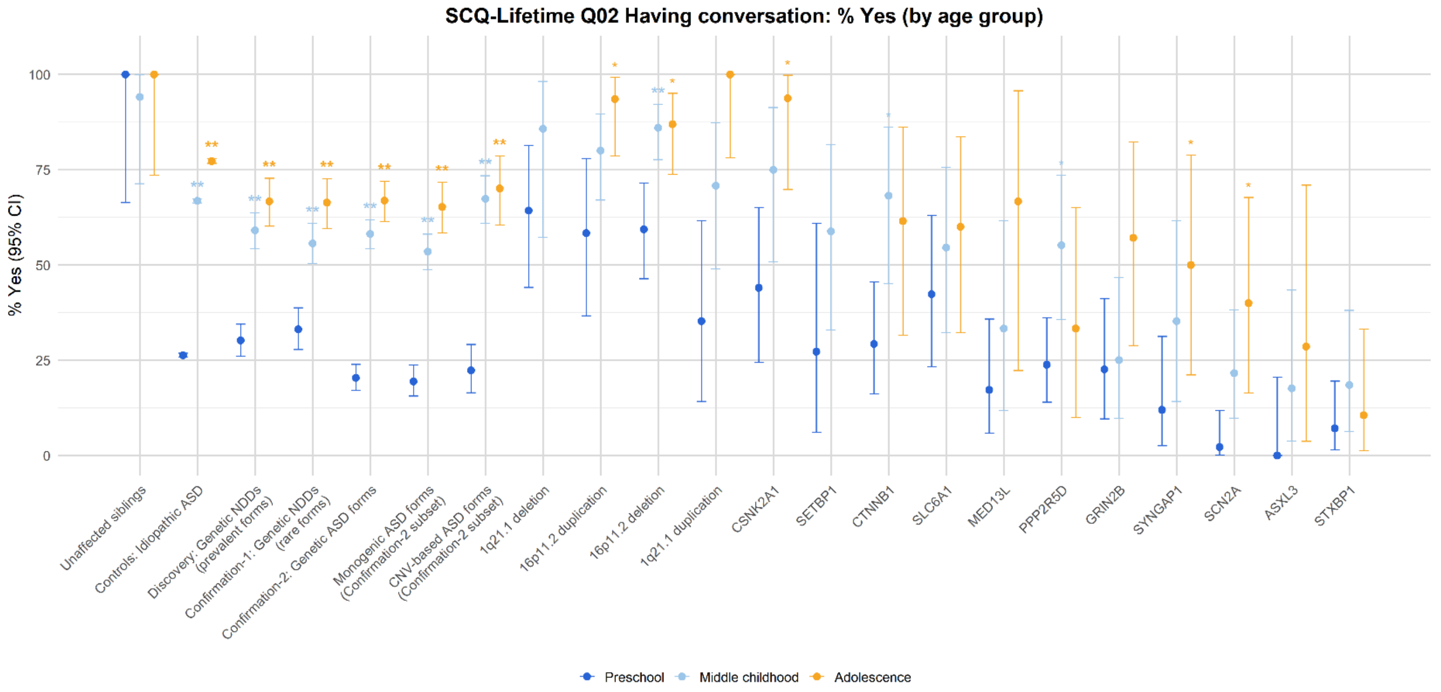


**Supplementary Figure 16: Age-stratified developmental trajectories for SCQ Q02 “Having conversation”** Shown are the group-level means and Clopper-Pearson 95% confidence intervals (CIs) of the SCQ Q02 “Having conversation” (percent “Yes”) outcome, stratified by age at evaluation: Preschool (0 to 5 years, 11 months; dark blue), Middle Childhood (6 to 11 years, 11 months; light blue), and Adolescence (12 to 17 years, 11 months; orange). Each age group is plotted separately for the 1) Unaffected siblings, 2) idiopathic ASD control, 3) *Discovery*, 4) *Confirmation-1*, and 5) *Confirmation-2* cohorts, followed by two subsets of the *Confirmation-2* cohort: 6) Monogenic ASD forms and 7) CNV-based ASD forms, and 8-22) 15 genetic conditions represented in the *Discovery* cohort. Within each genetic group, statistical comparisons were performed between Preschool (reference) and each older age group using binomial logistic regression with a logit link adjusted for sex and age at evaluation. Groups with fewer than five measurements were excluded from statistical testing and are not shown. Asterisks denote statistically significant group differences after Bonferroni multiple testing correction: * $P < 0.016$ (suggestive), ** $P < 7.94 \times 10^{-4}$ (significant).

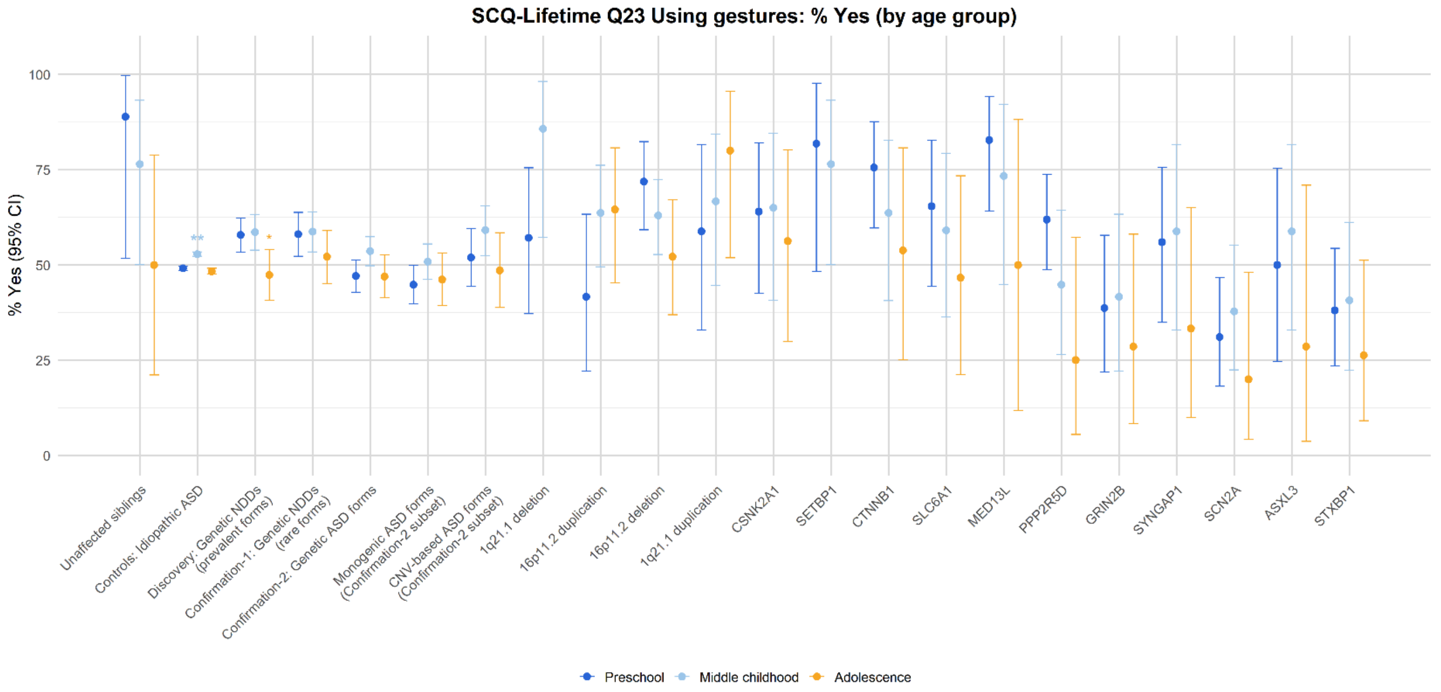


**Supplementary Figure 17: Age-stratified developmental trajectories for SCQ Q23 "Using gestures"**

Shown are the group-level means and Clopper-Pearson 95% confidence intervals (CIs) of the SCQ Q23 "Using gestures" (percent "Yes") outcome, stratified by age at evaluation: Preschool (0 to 5 years, 11 months; dark blue), Middle Childhood (6 to 11 years, 11 months; light blue), and Adolescence (12 to 17 years, 11 months; orange). Each age group is plotted separately for the 1) Unaffected siblings, 2) idiopathic ASD control, 3) *Discovery*, 4) *Confirmation-1*, and 5) *Confirmation-2* cohorts, followed by two subsets of the *Confirmation-2* cohort: 6) Monogenic ASD forms and 7) CNV-based ASD forms, and 8-22) 15 genetic conditions represented in the *Discovery* cohort. Within each genetic group, statistical comparisons were performed between Preschool (reference) and each older age group using binomial logistic regression with a logit link adjusted for sex and age at evaluation. Groups with fewer than five measurements were excluded from statistical testing and are not shown. Asterisks denote statistically significant group differences after Bonferroni multiple testing correction: * $P < 0.016$ (suggestive), ** $P < 7.94\times10^{-4}$ (significant).

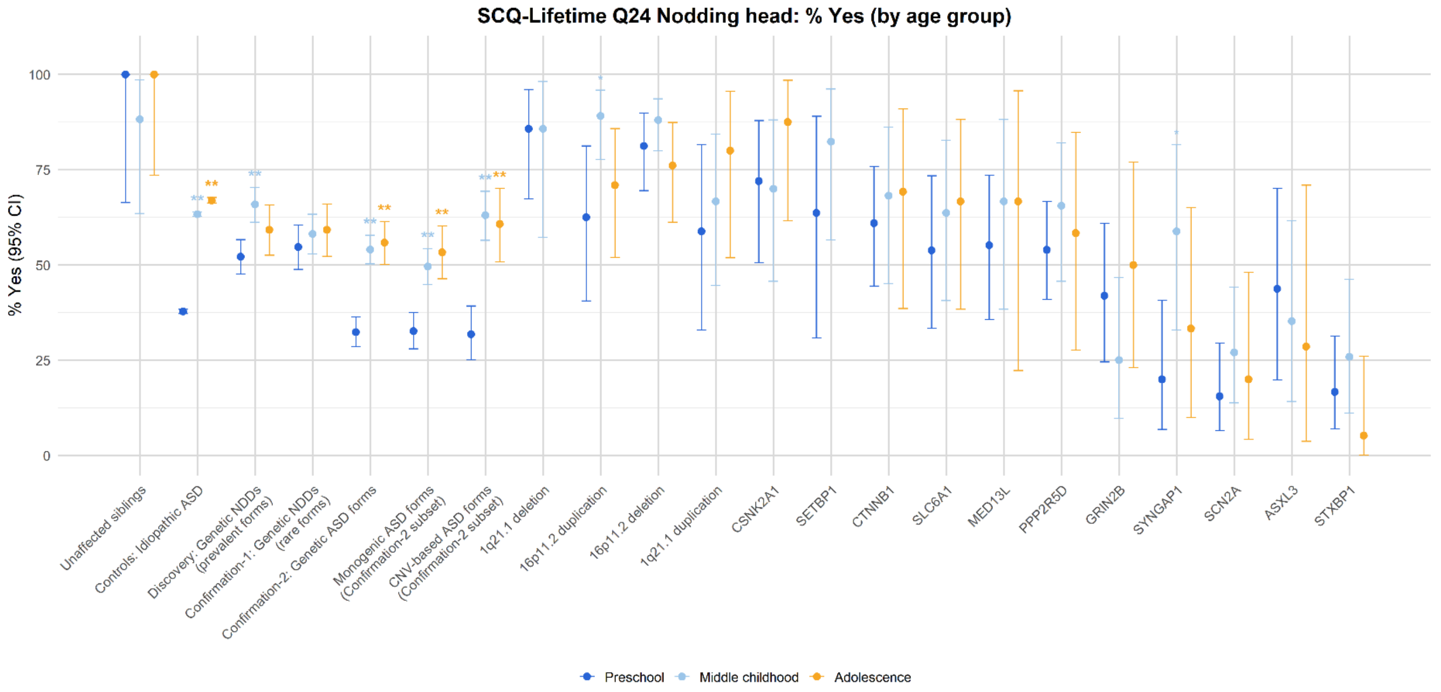


**Supplementary Figure 18: Age-stratified developmental trajectories for SCQ Q24 “Nodding head”**

Shown are the group-level means and Clopper-Pearson 95% confidence intervals (CIs) of the SCQ Q24 “Nodding head” (percent “Yes”) outcome, stratified by age at evaluation: Preschool (0 to 5 years, 11 months; dark blue), Middle Childhood (6 to 11 years, 11 months; light blue), and Adolescence (12 to 17 years, 11 months; orange). Each age group is plotted separately for the 1) Unaffected siblings, 2) idiopathic ASD control, 3) *Discovery*, 4) *Confirmation-1*, and 5) *Confirmation-2* cohorts, followed by two subsets of the *Confirmation-2* cohort: 6) Monogenic ASD forms and 7) CNV-based ASD forms, and 8-22) 15 genetic conditions represented in the *Discovery* cohort. Within each genetic group, statistical comparisons were performed between Preschool (reference) and each older age group using binomial logistic regression with a logit link adjusted for sex and age at evaluation. Groups with fewer than five measurements were excluded from statistical testing and are not shown. Asterisks denote statistically significant group differences after Bonferroni multiple testing correction: * $P < 0.016$ (suggestive), ** $P < 7.94\times10^{-4}$ (significant).

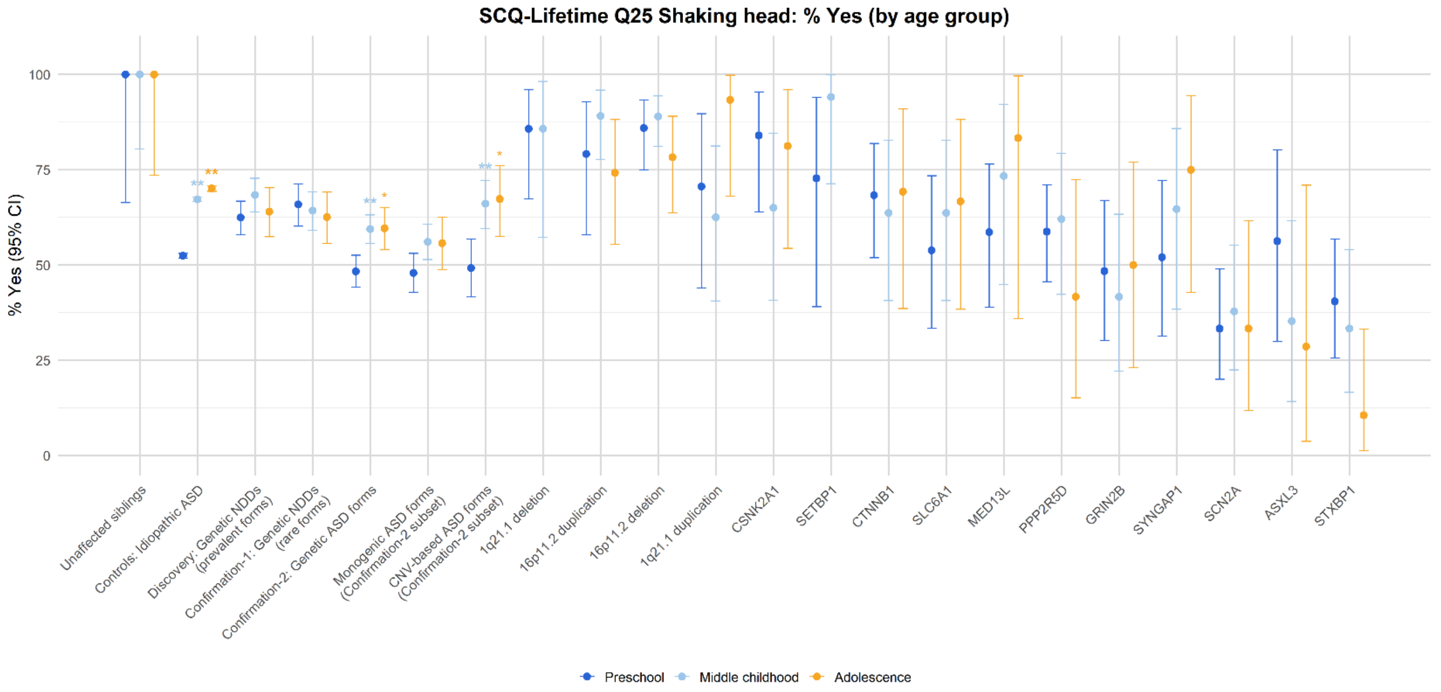


**Supplementary Figure 19: Age-stratified developmental trajectories for SCQ Q25 "Shaking head"**

Shown are the group-level means and Clopper-Pearson 95% confidence intervals (CIs) of the SCQ Q25 "Shaking head" (percent "Yes") outcome, stratified by age at evaluation: Preschool (0 to 5 years, 11 months; dark blue), Middle Childhood (6 to 11 years, 11 months; light blue), and Adolescence (12 to 17 years, 11 months; orange). Each age group is plotted separately for the 1) Unaffected siblings, 2) idiopathic ASD control, 3) *Discovery*, 4) *Confirmation-1*, and 5) *Confirmation-2* cohorts, followed by two subsets of the *Confirmation-2* cohort: 6) Monogenic ASD forms and 7) CNV-based ASD forms, and 8-22) 15 genetic conditions represented in the *Discovery* cohort. Within each genetic group, statistical comparisons were performed between Preschool (reference) and each older age group using binomial logistic regression with a logit link adjusted for sex and age at evaluation. Groups with fewer than five measurements were excluded from statistical testing and are not shown. Asterisks denote statistically significant group differences after Bonferroni multiple testing correction: * $P < 0.016$ (suggestive), ** $P < 7.94\times10^{-4}$ (significant).

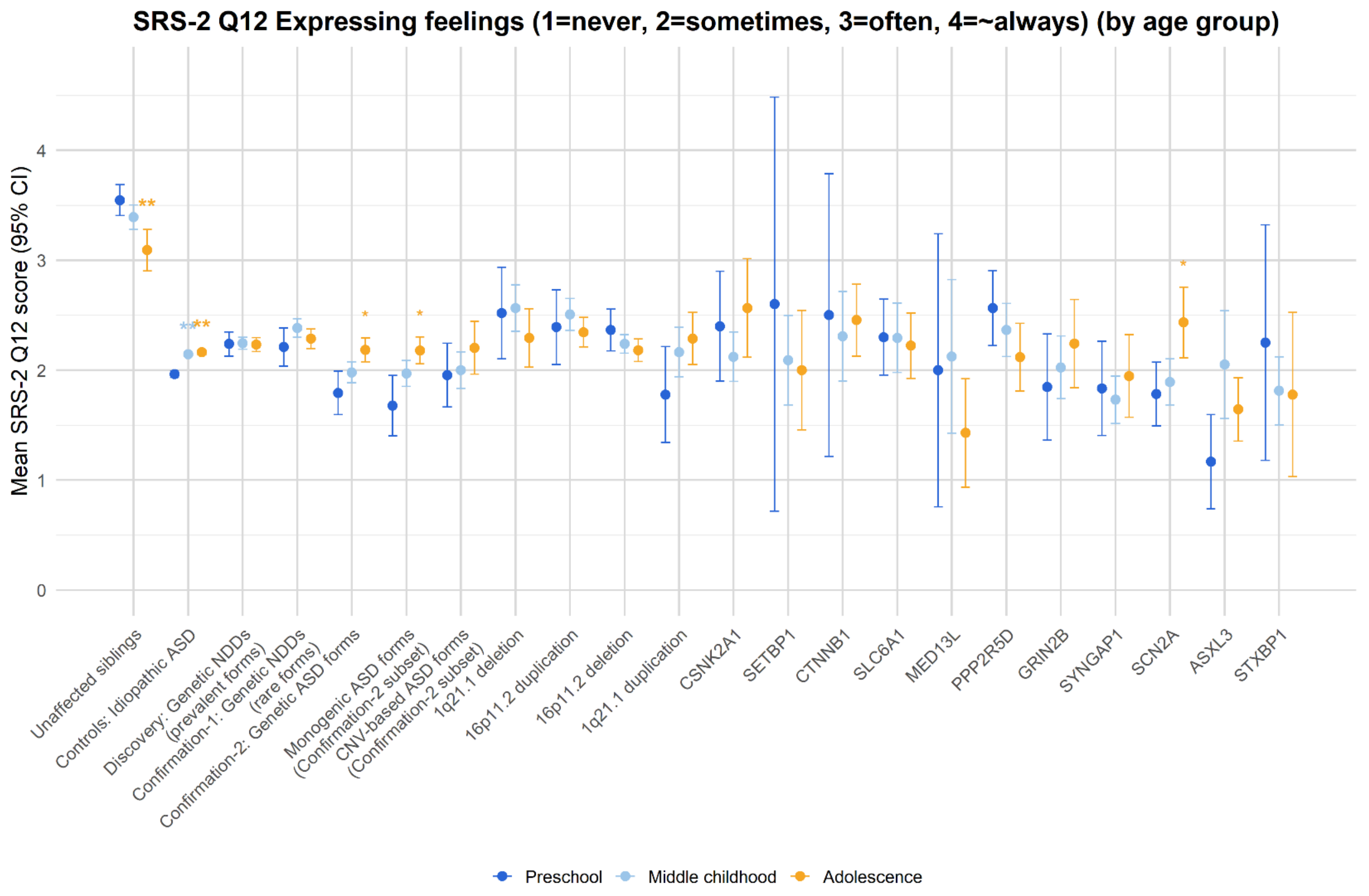


**Supplementary Figure 20: Age-stratified developmental trajectories for SRS-2 Q12 "Expressing feelings"** Shown are the group-level means and Clopper-Pearson 95% confidence intervals (CIs) of the SRS-2 Q12 "Expressing feelings" (ordinal: 1 = Not true; 2 = Sometimes true; 3 = Often true; 4 = Almost always true) outcome, stratified by age at evaluation: Preschool (0 to 5 years, 11 months; dark blue), Middle Childhood (6 to 11 years, 11 months; light blue), and Adolescence (12 to 17 years, 11 months; orange). Each age group is plotted separately for the 1) Unaffected siblings, 2) idiopathic ASD control, 3) *Discovery*, 4) *Confirmation-1*, and 5) *Confirmation-2* cohorts, followed by two subsets of the *Confirmation-2* cohort: 6) Monogenic ASD forms and 7) CNV-based ASD forms, and 8-22) 15 genetic conditions represented in the *Discovery* cohort. Within each genetic group, statistical comparisons were performed between Preschool (reference) and each older age group using binomial logistic regression with a logit link adjusted for sex and age at evaluation. Asterisks denote statistically significant group differences after Bonferroni multiple testing correction: * $P < 0.015$ (suggestive), ** $P < 7.58\times10^{-4}$ (significant).
